# Phase II/III Double-Blind Study Evaluating Safety and Immunogenicity of a Single Intramuscular Booster Dose of the Recombinant SARS-CoV-2 Vaccine “Patria” (AVX/COVID-12) Using an Active Newcastle Disease Viral Vector (NDV) during the Omicron Outbreak in Healthy Adults with Elevated Baseline Antibody Titers from Prior COVID-19 and/or SARS-CoV-2 Vaccination

**DOI:** 10.1101/2024.02.11.24302530

**Authors:** Constantino López-Macías, Martha Torres, Brenda Armenta-Copca, Niels H. Wacher, Arturo Galindo-Fraga, Laura Castro-Castrezana, Andrea Alicia Colli-Domínguez, Edgar Cervantes-Trujano, Isabel Erika Rucker-Joerg, Fernando Lozano-Patiño, Juan José Rivera-Alcocer, Abraham Simón-Campos, Efrén Alberto Sánchez-Campos, Rafael Aguirre-Rivero, Alejandro José Muñiz-Carvajal, Luis del Carpio-Orantes, Francisco Márquez-Díaz, Tania Rivera-Hernández, Alejandro Torres-Flores, Luis Ramírez-Martínez, Georgina Paz-De la Rosa, Oscar Rojas-Martínez, Alejandro Suárez-Martínez, Gustavo Peralta-Sánchez, Claudia Carranza, Esmeralda Juárez, Horacio Zamudio-Meza, Laura E. Carreto-Binaghi, Mercedes Viettri, Damaris Romero-Rodríguez, Andrea Palencia, David Sarfati-Mizrahi, Weina Sun, Héctor Elías Chagoya-Cortés, Felipa Castro-Peralta, Peter Palese, Florian Krammer, Adolfo García-Sastre, Bernardo Lozano-Dubernard

**Author notes:** Correspondence (B. Lozano-Dubernard), (C. López-Macías).

## Abstract

**Background:** The urgent need for safe, effective, and economical coronavirus disease 2019 (COVID-19) vaccines, especially for booster campaigns targeting vulnerable populations, prompted the development of the AVX/COVID-12 vaccine candidate. AVX/COVD-12 is based in a Newcastle disease virus La Sota (NDV-LaSota) recombinant viral vector. This vaccine expresses a stabilized version of the spike protein of severe acute respiratory syndrome coronavirus 2 (SARS-CoV-2), specifically the ancestral Wuhan strain. The study aimed to assess its safety, immunogenicity, and potential efficacy as an anti-COVID-19 booster vaccine.

**Methods:** In a phase II/III clinical trial conducted from November 9, 2022, to September 11, 2023, a total of 4,056 volunteers were enrolled. Participants received an intramuscular booster dose of either AVX/COVID-12 or AZ/ChAdOx-1-S vaccines. Safety, immunogenicity, and potential efficacy were assessed through various measures, including neutralizing antibody titers, interferon (IFN)-γ-producing CD4+ T cells, and CD8+ T cells. The evaluation also involved immunobridging, utilizing the AZ/ChAdOx-1-S vaccine as an active comparator, and monitoring the incidence of COVID-19 cases.

**Findings:** The AVX/COVID-12 vaccine induced neutralizing antibodies against both the ancestral SARS-CoV-2 and the BA.2 and BA.5 Omicron variants. The geometric mean ratio of neutralizing antibody titers between individuals immunized with the AVX/COVID-12 vaccine and those with the AZ/ChAdOx-1-S vaccine at 14 days is 0.96, with a confidence interval (CI) of 0.85-1.06. The outcome aligns with the non-inferiority criterion recommended by the World Health Organization (WHO), indicating a lower limit of the CI greater than or equal to 0.67. Induction of IFN-γ-producing CD8+ T cells at day 14 post-immunization was exclusively observed in the AVX/COVID-12 group. Finally, a trend suggested a potentially lower incidence of COVID-19 cases in AVX/COVID-12 boosted volunteers compared to AZ/ChAdOx-1-S recipients.

**Conclusion:** The AVX/COVID-12 vaccine proved safe, well-tolerated, and immunogenic. AVX/COVID-12 meets the WHO non-inferiority standard compared to AZ/ChAdOx-1-S. These results strongly advocate for AVX/COVID-12 as a viable booster dose, supporting its utilization in the population.

## Introduction

Severe acute respiratory syndrome coronavirus 2 (SARS-CoV-2) and its variants of concern have caused hundreds of millions of infections and millions of deaths around the globe since 2020 (1).

At the beginning of 2021, anti-SARS-CoV-2 vaccination campaigns had been launched in almost 200 countries with more than 13.53 billion doses distributed (2); as of November 2022, emergency use approval has been granted to 50 vaccine candidates (3). These vaccines have shown to be effective in phase III clinical trials and to induce neutralizing antibodies that target the spike protein of SARS-CoV-2 among other antigens less studied (4). In addition to the vaccines already in use, many other candidates are in a preclinical or clinical development.

Despite the remarkable speed in the development of coronavirus disase 2019 (COVID-19) vaccines since 2020, there have been over 6.9 million official deaths from COVID-19 (1) (with estimates based on excess mortality being 3-4 times higher). Moreover, the unequal global distribution of vaccines, particularly impacting low-and middle-income countries (LMICs), underscores the urgency for the continued development of new vaccine platforms to address disparities in distribution. Ensuring a high vaccination rate worldwide is crucial for reducing the likelihood of new variant emergence. Additionally, the administration of periodic booster doses is essential to prevent subsequent waves of COVID-19 and safeguard vulnerable populations, including the elderly, immunocompromised individuals, and those with comorbidities. Consequently, the global vaccine supply remains an urgent issue that requires attention (5).

The AVX/COVID-12 vaccine candidate is a Newcastle disease virus LaSota (NDV-LaSota) vector-based vaccine that expresses the HexaPro version (6 prolines that stabilize the prefusion conformation) of the SARS-CoV-2 spike protein (6,7).

Among the main advantages offered by the NDV-LaSota vector is the standardized production process in embryonated chicken eggs, easy to develop in existing facilities for the manufacture of influenza vaccines, achieving the production of large numbers of doses in a short time (8). Therefore, this could help to fulfill vaccine demand, particularly in countries with a low immunization rate or incomplete vaccination scheme.

The NDV-LaSota vector, expressing the spike protein of the SARS-CoV-2 virus, has demonstrated efficacy in preclinical studies, particularly in mice and hamsters. Following intranasal (IN) immunization in mice, the generation of IgA and IgG2a antibodies, with a response of IFN-γ-producing T cells, was observed. This vector provided complete protection against infection in mice and hamsters, indicated by the absence of viral titers and viral antigen in the lungs of mice and a significant reduction in SARS-CoV-2 shedding in hamsters (8,9).

Preclinical trials involving pigs were conducted in Mexico to assess the safety and immunogenicity of the AVX/COVID-12 vaccine when administered via intramuscular (IM) or IN routes. The study revealed robust responses in serum-neutralizing antibodies, with notable reactivity to variants of concern (VOCs) carrying mutations in the spike protein epitopes (10). In a Phase I clinical trial (NCT04871737) with 91 volunteers, the vaccine was evaluated with various booster regimens administered through both IM and IN routes, demonstrating significant immunogenicity in both cases. The safety of this vaccine was also confirmed in these studies, establishing the foundation for further clinical development (11).

Similar vaccines are undergoing clinical trials, with a live version being tested in the United States (NCT05181709), whereas inactivated vaccines based on the same vector are also in clinical testing, with ongoing trials in Vietnam (Phase I NCT04830800), Thailand (Phase I/II NCT04764422, Phase III TCTR20221026004), and Brazil (Phase I NCT04993209 and Phase II/III NCT05354024). Available reports indicate favorable safety and immunogenicity profiles for these vaccines (12–14). Recently, an inactivated version of this vaccine was authorized for emergency use as a booster for the prevention of severe COVID-19 in Thailand (15).

In Phase II clinical trials, the AVX/COVID-12 vaccine underwent assessment through both IM and IN administrations as a booster dose. The study demonstrated safety and immunogenicity via both routes in volunteers who had previously received mRNA, AZ/ChAdOx-1-S, or inactivated virus vaccines or were infected with SARS-CoV-2, all of whom initially exhibited anti-spike IgG titers below 1,200 units (U)/mL in a chemiluminescence test. AVX/COVID-12 IM or IN immunization induced a significant (≥ 2.5 fold) increase in the neutralizing antibody response against SARS-CoV-2 variants of concern by day 14, despite expressing the original Wuhan-1 strain spike protein. Noteworthy elevations in neutralizing antibodies against multiple variants of concern, including Alpha, Beta, Delta, and Omicron (BA.2 and BA.5), were observed when compared to the placebo groups (16).

In this phase II/III parallel study, we investigated the safety and immunogenicity of a single intramuscular booster dose of AVX/COVID-12 in healthy adults with a history of prior COVID-19 vaccination and/or SARS-CoV-2 infection. Unlike in the Phase II trial, participants were included irrespective of their initial anti-S antibody levels. Given the pandemic situation during the recruiting period of the trial (Figure 1S), enrolled individuals exhibited a wide range of baseline antibody titers. Additionally, we conducted an active-controlled non-inferiority study (immunobridging) using the AZ/ChAdOx-1-S vaccine as the comparator, as both are vectorized vaccines. The AZ/ChAdOx-1-S vaccine has proven to be safe and effective against SARS-CoV-2 infection, demonstrating an effectiveness of 84.3% against asymptomatic SARS-CoV-2 infection and 98.9% against COVID-19 hospitalization (17,18). Moreover, it has become the most widely used vaccine globally, deployed in 185 countries since its approval for emergency use (19). In Mexico, since November 2021, more than 79.43 million doses have been administered, covering single-dose, full vaccination, and booster regimens, utilizing both homologous and heterologous systems (20). The characteristics of AZ/ChAdOx-1-S make it a suitable comparator for testing AVX/COVID-12 in a non-inferiority trial.

**Figure 1.**
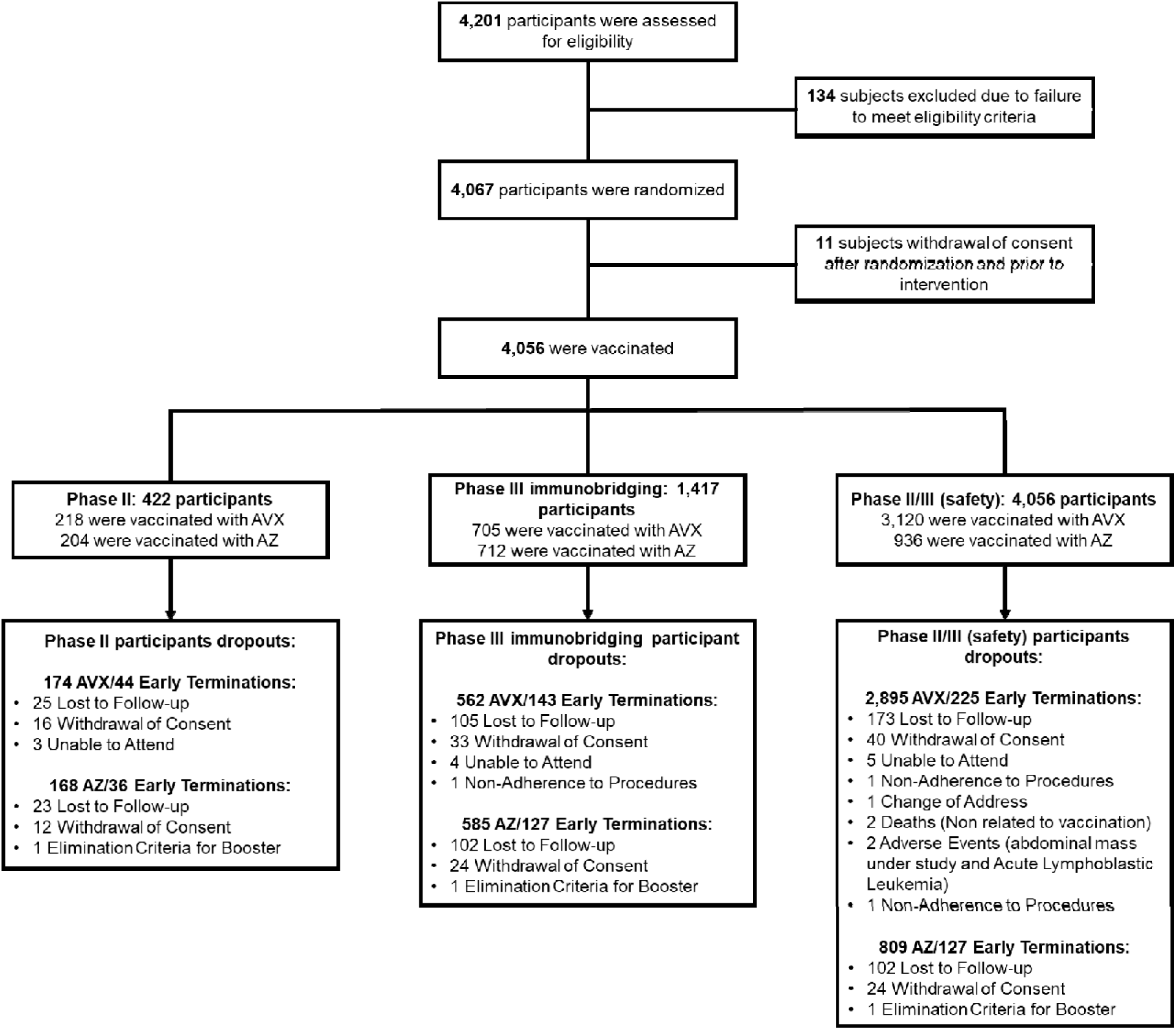
Participant Screening and Randomization. A flowchart illustrating the screening, randomization, and distribution of participants among different experimental groups throughout the study.

## Material and methods

### Study design

A phase II/III double-blind study (ClinicalTrials.gov #NCT05710783) was conducted to assess the safety, immunogenicity, and post-boosting incidence of SARS-CoV-2 infection following a single IM dose of SARS-CoV-2 vaccine (AVX/COVID-12) “Patria”. The primary objective was to establish non-inferiority in the SARS-CoV-2 neutralizing antibodies production at day 14 post-administration by comparing it with an active control, the AZ/ChAdOx-1-S vaccine. Randomization involved the administration of a single IM dose of the experimental AVX/COVID-12 vaccine at a concentration of 10^8.0^ tissue culture infective dose 50% (TCID50%) per dose and a single IM dose of the AZ/ChAdOx-1-S vaccine at a concentration of 10^10^ viral particles per dose for subsequent non-inferiority analysis.

The protocol was formulated by iLS Clinical Research, S.C. in collaboration with the Mexican Institute for Social Security (IMSS) and Laboratorio Avi-Mex, S.A. de C.V. (Avimex®), which was the sponsor. Approval for the study was granted by the Federal Commission for the Protection against Sanitary Risks (COFEPRIS) in Mexico, assigned the number RNEC2022-AVXSARSCoV2VAC005. Ethics approval was obtained from the institutional ethics committee at each research site participating in the study. The research was conducted in complete compliance with Mexican regulations and in accordance with the principles outlined in the Declaration of Helsinki and Good Clinical Practice. Immunological assay samples were processed at the National Institute for Respiratory Diseases (INER) in Mexico City.

### Study groups

The study groups comprised adults aged 18 years or older, of both genders, who were in good health and had received at least one dose of an approved SARS-CoV-2 vaccine. Inclusion criteria for the study involved a negative PCR test for SARS-CoV-2 during the screening visit, a negative pregnancy test in women of childbearing potential, a commitment to maintaining adequate prevention measures to avoid infection with SARS-CoV-2 throughout their participation in the study, especially during the first 14 days after the baseline visit. Importantly, there were no restrictions based on the baseline antibody titers for eligibility in the study.

The exclusion criteria encompassed hypersensitivity or allergy to any ingredient of the vaccine, a history of severe anaphylactic reactions, fever at the baseline visit, receipt of the last COVID-19 vaccination within the past 4 months, SARS-CoV-2 infection within the last month, pregnancy or nursing status, chronic diseases requiring the use of immunosuppressive agents or immune response modulators (e.g., systemic corticosteroids, cyclosporine, rituximab), active chemotherapy treatment for cancer, a history of human immunodeficiency virus (HIV) infection, and chronic renal or liver disease with a recent history of infectious conditions requiring hospitalization or intravenous drug treatment within the year before the baseline visit.

The randomization of volunteers was conducted using a computer-based assignment system, ensuring blinding throughout the study. Upon signing the informed consent form, each participant received a patient number, encoding all their information in a pseudo-anonymized manner during data collection and becoming fully anonymized during the analysis. The stratification for subject randomization is as follows: In phase 2 and the immunobridging phase, blocks of 8 were created, with each block randomly assigning 4 subjects to receive the AVX/COVID-12 vaccine and 4 subjects to receive the AZ/ChAdOx-1-S vaccine. In the follow-up safety phase III, blocks of 8 were also created, with all subjects receiving the AVX/COVID-12 vaccine.

In phase II, participants were enrolled to receive either the AVX/COVID-12 or AZ/ChAdOx-1-S vaccine. During phase III immunobridging, the phase II participants were included, and new participants were enrolled. For the phase II/III safety assessment, volunteers from the phase III immunobridging and phase II participants were included. Additionally, more volunteers were recruited to receive the AVX/COVID-12 vaccine (Figure 1).

### Safety

Safety was assessed through active and passive monitoring of adverse events (AEs) in both the AZ/ChAdOx-1-S and AVX/COVID-12 vaccine groups, taking into account their severity and their relationship to the intervention. The study comprised four visits, with the option for additional visits in case of suspected SARS-CoV-2 infection. While all participants completed the same visits, those assigned to different groups (described above and in Figure 1) underwent additional sample collections during visits on days 0, 14, 90, and 180 of the study.

The primary endpoints in the phase II/III and the immunobridging aspects of this trial included solicited local and systemic reactions. These reactions were assessed at the time participants entered the study and continued throughout a 6-month follow-up period. Additionally, the study reported the incidence of symptomatic, severe, or fatal cases of COVID-19.

The classification of potential AEs and their association with vaccine exposure followed a two-period analysis. The first period included the initial 7 days following vaccination, during which adverse reactions were deemed related to the vaccine components (19). Conversely, reactions occurring after 7 days, typically in response to the development of an immune response, constituted the second analysis period (19).

### Sample collection

Venous blood samples were obtained from participants during scheduled site visits for serology tests and cellular response assessments. The samples were collected into two sodium heparin tubes and one separator tube (SST BD vacutainer tubes, Franklin Lakes, NJ, USA) through standard phlebotomy procedures. Following collection, the samples were transported at room temperature to the INER for immediate processing. All blood samples and their products were handled in a biosafety level (BSL)-2 laboratory, employing appropriate personal protective equipment and safety precautions, adhering to processing protocols approved by the Institutional Biosafety Committee. Serum isolation was performed by centrifuging venous blood collected into SST at 2,260 x g for 10 minutes to separate the serum. The resulting serum was carefully extracted from the upper portion of the tube, aliquoted, and then stored at −20 °C until needed.

### Peripheral blood mononuclear cell (PBMC) isolation

PBMC were isolated from venous blood collected in sodium heparin tubes (BD vacutainer tubes, Franklin Lakes, NJ, USA). Within four hours of collection, PBMC isolation was conducted by density-gradient sedimentation of whole blood diluted at a 1:2 ratio in Roswell Park Memorial Institute (RPMI) 1640 medium (Lonza, Basel, Switzerland) at room temperature. The diluted blood was layered over an appropriate volume of room-temperature Lymphoprep (Axis-Shield Diagnostics, Dundee, UK). Then, the PBMC were then recovered, cryopreserved in a medium consisting of 10% dimethyl sulfoxide (DMSO) (Sigma Aldrich, St. Louis, MO, USA) and 90% heat-inactivated fetal bovine serum (FBS; GIBCO, California, USA), and stored at −80 °C until use. For T cell assays, the thawing process involved warming frozen cryovials at 37 °C in a water bath, and cells were resuspended in 5 mL of CTS OpTmizer (GIBCO, California, USA). After a wash with OpTmizer, cells were counted using a Neubauer chamber, and the viability was assessed by trypan blue. Cells were resuspended in OpTmizer medium to a density of 4 x 10^6^ cells/mL.

The subunit 1 of the spike protein of SARS-CoV-2 (RayBiotech, Peachtree Corners, GA, USA) was selected as the primary antigen due to its display of most immunogenic epitopes of the spike protein. Additionally, it contains the receptor binding domain (RBD) fragment of the virus, which mediates binding to the host receptor angiotensin-converting enzyme 2 (ACE2).

### Pseudovirus neutralization assay

The pseudovirus neutralization assay employed pseudovirus particles based on replication-competent vesicular stomatitis virus (VSV)-eGFP-SARS-CoV-2. These particles encoded the spike gene of the ancestral Wuhan-1 strain, as well as Omicron subvariants BA.2 (VSV-SARS-CoV-2 BA.2 clone 1) and BA.5 (VSV-SARS-CoV-2 BA.5 clone 4). The pseudovirus particles were generously provided by Sean Whelan and used in pseudoviral microneutralization assays.

Prior to use in the neutralization assay, all sera were heat-inactivated at 56 °C for 30 minutes. Vero E6 cells (ATCC CRL-1586, Manassas, VA, USA) were seeded at a density of 1.3 x 10^4^ cells per well in 96-well plates with 100 µL of complete EMEM (Eagle’s minimum essential medium, ATCC, 30-2003 Manassas, VA, USA) supplemented with 10% fetal bovine serum; (FBS; Gibco, Waltham, MA, USA). The cells were then incubated for 24 hours at 37 °C with 5% CO_2_, allowing them to reach approximately 85% confluence.

The next day, serum samples were diluted in EMEM medium without FBS using a 3-fold serial dilution, starting with a 1:60 dilution. These diluted samples were mixed with 100 TCID_50_ of VSV-SARS-CoV-2 spike pseudoparticles of the ancestral Wuhan-1 strain or Omicron variants. The mixture was incubated for 60 minutes at 37 °C with 5% CO_2_ to facilitate the neutralization process. Subsequently, each dilution was transferred to a parallel plate containing Vero E6 monolayers and incubated at 37°C with 5% CO_2_ for 72 hours.

After the incubation period, 3.7% formaldehyde was added to the culture plates and the plates were incubated 30-40 minutes at room-temperature. Then, the formaldehyde was discarded, and the plates were washed twice with phosphate-buffered saline (PBS; Lonza, Basel, Switzerland). The neutralizing activity of each sample was reported as the titer, represented by the reciprocal of dilution, where the cell protection is 50%; to get this value, we considered the last serum dilution at which the wells are non-infected and with this data, the half maximal 50% inhibitory dilution (ID_50_) was calculated using the Spearman-Kaerber analysis method. This analysis was based on the presence or absence of cellular damage (cytopathic effect). Each sample was tested in duplicate.

### Intracellular cytokine staining assay

Cells were costimulated for 16 hours with costimulation anti-human CD28/CD49d (BD Biosciences, San Jose, CA, USA), spike subunit 1 protein at a concentration of 5 µg/mL. Medium served as a negative control, and phytohemagglutinin (PHA; 1 mg/mL) as a positive control. Monensin (GolgiStop BD, San José, CA, USA) was added, and the samples were incubated for 4 hours at 37 °C.

After stimulation, the samples were washed with PBS (Lonza, Walkersville, MD, USA) and stained with Live/Dead Fixable near 633 or 635nm (Invitrogen, Waltham, MA, USA) (diluted 1:1000) for 15 minutes at room temperature in the dark. Subsequently, cell surface staining was performed using a cocktail of anti-human CD3-Alexa Fluor 700 (Biolegend, San Diego, CA, USA), CD4-PerCP/Cy5.5 (Biolegend, San Diego, CA, USA), and CD8-PE/Cy7 (Biolegend, San Diego, CA, USA) antibodies in cell staining buffer (BioLegend, San Diego, CA, USA). The mixture was incubated for 15 minutes at room temperature in the dark, followed by an additional washing step with cell staining buffer.

The cells were fixed and permeabilized using Cytofix/Cytoperm (BD, San Jose, USA) for 20 minutes at room-temperature. After centrifugation, Perm/Wash (BD, New Jersey, USA) was added. Then, intracellular staining was then performed using a cocktail of anti-human IFN-γ-FITC (BD, San Jose, USA), TNF-α APC (Biolegend, San Diego, CA, USA), and IL-2-BV605 (Biolegend, San Diego, CA, USA) antibodies in Perm/Wash solution for 30 minutes at room temperature in the dark. After washing with BD Perm/Wash buffer (BD, San Jose, USA), the cells were resuspended in PBS and kept at 4 °C in the dark until acquisition and analysis. Unstained and fluorescence minus one (FMO) controls were included.

Sample acquisition was carried out on a BD Symphony A1 instrument, previously calibrated with standardized Cytometer Setup and Tracking (CST) beads (BD, San Jose, USA) used for daily quality control. Compensation was performed using CompBeads (BD, San Jose, USA). At least 200,000 events of the lymphocyte region in a Forward Scatter (FSC) vs Side Scatter (SSC) plot were acquired per sample. Analysis was conducted using FACSDiva 8.0.3, and gates for identification of SARS-CoV-2 antigen-specific cytokine-producing CD3+, CD4+, or CD8+ T cells were defined using the FMO controls. This study was approved by the Institutional Review Board for Human Subjects Research at INER.

### Statistical analysis

We present results encompassing safety, immunogenicity, efficacy, and a sample size determined through statistical hypothesis testing. To outline the study population demographic characteristics, measures of central tendency and dispersion were employed for continuous variables, while proportions were utilized for discrete or categorical variables. Continuous variables underwent analysis using the student’s t-test or non-parametric test where specified, and the categorical variables were compared through a chi-square test.

Safety analysis involved the AEs comparison in participants vaccinated with AZ/ChAdOx-1-S (control group) and AVX/COVID-12 (experimental group), considering both severity and their relationship to the intervention. The Z-test was employed to compare the proportions of reported adverse effects between treatment groups.

Variations in the neutralizing antibody titers and secreted IFN-γ levels were determined. Statistically significant differences in intra-groups in geometric mean neutralization titers between day 0 and day 14, 90, and 180 were analyzed using a paired t-test for the variable on a logarithmic scale, and the differences in IFN-γ production were determined using Wilcoxon signed ranks test. Significance was set at p-values < 0.05. The confidence interval for the non-inferiority criteria was obtained through a student’s t-test for the variable on a logarithmic scale.

The analysis of the incidence of symptomatic, severe, or fatal cases of COVID-19 is described by Nelson Alen’s cumulative hazard rate, and the difference between groups was evaluated with a log rank test.

The statistical analysis was performed with STATA v 17.0 (StataCorp, College Station, TX, USA).

## Results

### Randomization of study participants

From November 9, 2022, to September 11, 2023, a total of 4,201 volunteers underwent eligibility assessments, and 134 subjects were excluded due to failure to meet eligibility criteria. The distribution of recruited volunteers is depicted in Figure 1. Various research sites across different states of Mexico participated in volunteer recruitment, including Aguascalientes (Hospital Cardiológica Aguascalientes), Guerrero (CICPA Centro de Investigación Clínica del Pacífico), Mexico City (CAIMED Investigación en Salud, S.A. de C.V., Centro de Investigación Clínica Chapultepec, CICA Centro de Investigación Clínica Acelerada, S.C., INCMNSZ Instituto Nacional de Ciencias Médicas y Nutrición Salvador Zubirán, Unidad de Medicina Familiar No. 20 IMSS), Michoacán (Sociedad Administradora de Servicios de Salud, S.C.), Oaxaca (OSMO Oaxaca Site Management Organization S.C.), Quintana Roo (Red OSMO Centro de Investigación y Avances Médicos Especializados), State of Mexico (CRI Clinical Research Institute S.C.), Veracruz (Instituto Veracruzano de Investigación Clínica S.C.), and Yucatán (Centro Multidisciplinario para el Desarrollo Especializado de la Investigación Clínica en Yucatán (CEMDEICY) S.C.P., K&M Köhler & Milstein Facultad de Medicina, Universidad Autónoma de Yucatán, Mérida, Unidad de Atención Médica e Investigación en Salud (UNAMIS)).

A total of 4,067 participants were initially randomized for the study, with 11 withdrawing consent post-randomization, and 4,056 were vaccinated. In phase II, 422 participants were enrolled, with 218 receiving the AVX/COVID-12 vaccine and 204 administered the AZ/ChAdOx-1-S vaccine. In phase III immunobridging, 1,417 participants were enrolled, including 705 who received the AVX/COVID-12 vaccine and 712 who received the AZ/ChAdOx-1-S vaccine. For phase II, 44 AVX/COVID-12 and 36 AZ/ChAdOx-1-S vaccinees dropped out, leaving 174 AVX/COVID-12 and 168 AZ/ChAdOx-1-S participants who completed the study. In phase III immunobridging, 143 AVX/COVID-12 and 127 AZ/ChAdOx-1-S vaccinees dropped out, leaving 562 AVX/COVID-12 and 585 AZ/ChAdOx-1-S participants who completed the study. For phase II/III safety, 4,056 participants were enrolled, 3,120 received AVX/COVID-12. In addition, 936 volunteers were vaccinated with AZ/ChAdOx-1-S. 225 dropouts were recorded from the AVX/COVID-12 group and 127 from the AZ/ChAdOx-1-S group (Figure 1).

For the groups vaccinated with AVX/COVID-12 and AZ/ChAdOx-1-S, the average age was 39 years. No statistically significant differences were observed between the groups in terms of age, sex, race, or comorbidities, suggesting that both groups are comparable concerning these variables (Table 1).

**Table 1.**
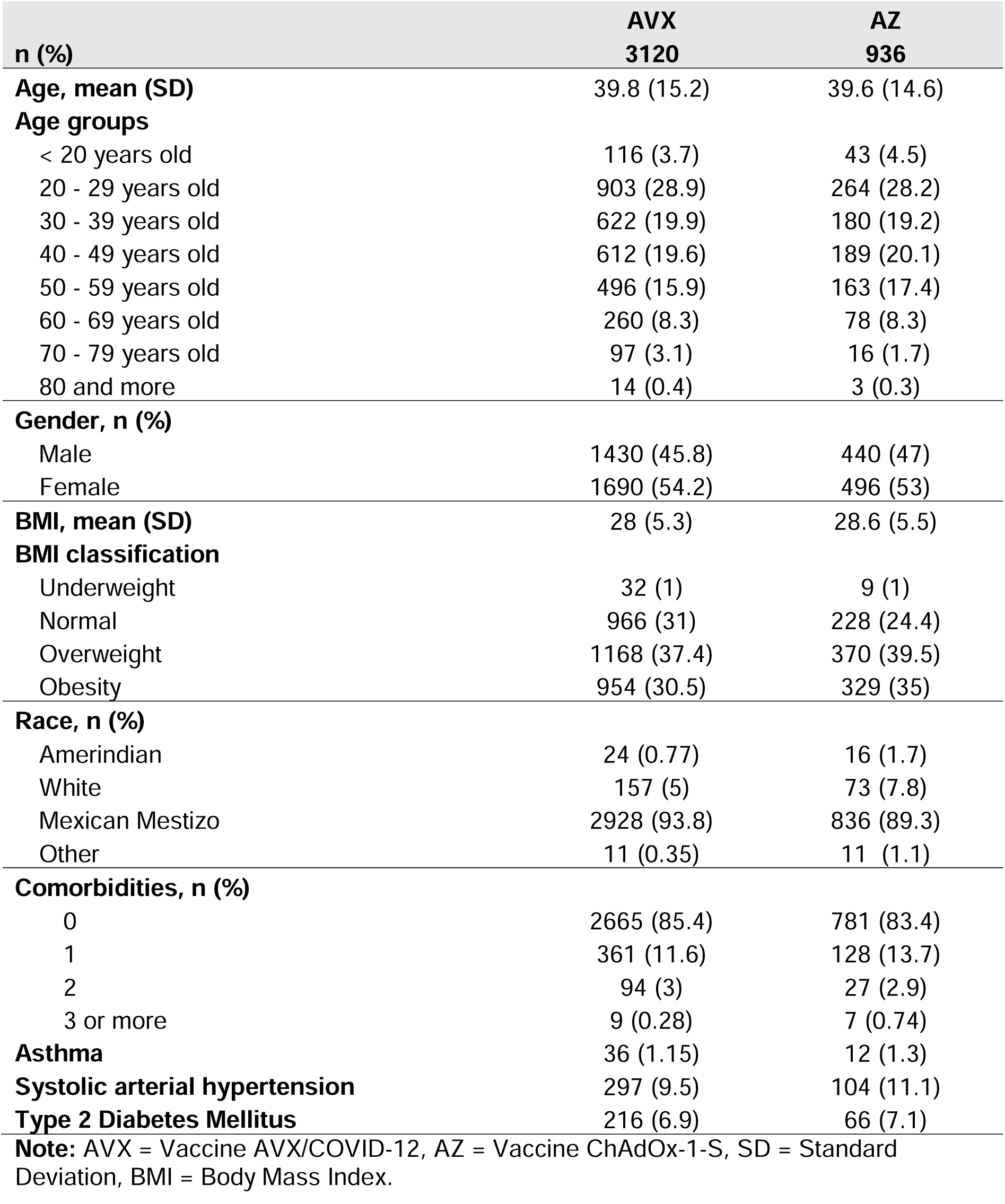
Phase II/III demographic characteristics.

| **n (%)** | **AVX**<br>**3120** | **AZ**<br>**936** |
| --- | --- | --- |
| **Age, mean (SD)** | 39.8 (15.2) | 39.6 (14.6) |
| **Age groups** |  |  |
| < 20 years old | 116 (3.7) | 43 (4.5) |
| 20 - 29 years old | 903 (28.9) | 264 (28.2) |
| 30 - 39 years old | 622 (19.9) | 180 (19.2) |
| 40 - 49 years old | 612 (19.6) | 189 (20.1) |
| 50 - 59 years old | 496 (15.9) | 163 (17.4) |
| 60 - 69 years old | 260 (8.3) | 78 (8.3) |
| 70 - 79 years old | 97 (3.1) | 16 (1.7) |
| 80 and more | 14 (0.4) | 3 (0.3) |
| **Gender, n (%)** |  |  |
| Male | 1430 (45.8) | 440 (47) |
| Female | 1690 (54.2) | 496 (53) |
| **BMI, mean (SD)** | 28 (5.3) | 28.6 (5.5) |
| **BMI classification** |  |  |
| Underweight | 32 (1) | 9 (1) |
| Normal | 966 (31) | 228 (24.4) |
| Overweight | 1168 (37.4) | 370 (39.5) |
| Obesity | 954 (30.5) | 329 (35) |
| **Race, n (%)** |  |  |
| Amerindian | 24 (0.77) | 16 (1.7) |
| White | 157 (5) | 73 (7.8) |
| Mexican Mestizo | 2928 (93.8) | 836 (89.3) |
| Other | 11 (0.35) | 11 (1.1) |
| **Comorbidities, n (%)** |  |  |
| 0 | 2665 (85.4) | 781 (83.4) |
| 1 | 361 (11.6) | 128 (13.7) |
| 2 | 94 (3) | 27 (2.9) |
| 3 or more | 9 (0.28) | 7 (0.74) |
| **Asthma** | 36 (1.15) | 12 (1.3) |
| **Systolic arterial hypertension** | 297 (9.5) | 104 (11.1) |
| **Type 2 Diabetes Mellitus** | 216 (6.9) | 66 (7.1) |

**Note:** AVX = Vaccine AVX/COVID-12, AZ = Vaccine ChAdOx-1-S, SD = Standard Deviation, BMI = Body Mass Index.

Information regarding prior SARS-CoV-2 infection and vaccination history was collected from participants in the immunobridging study (Table 2). Results show no statistically significant difference in the average time since the last infection and that the proportion of subjects with a history of SARS-CoV-2 infection is the same for both groups. Similarly, the proportion of subjects vaccinated with different vaccine technologies is comparable, considering the number of doses received.

**Table 2.**
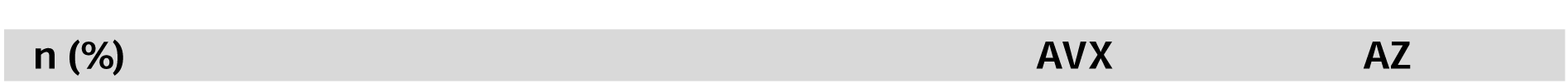
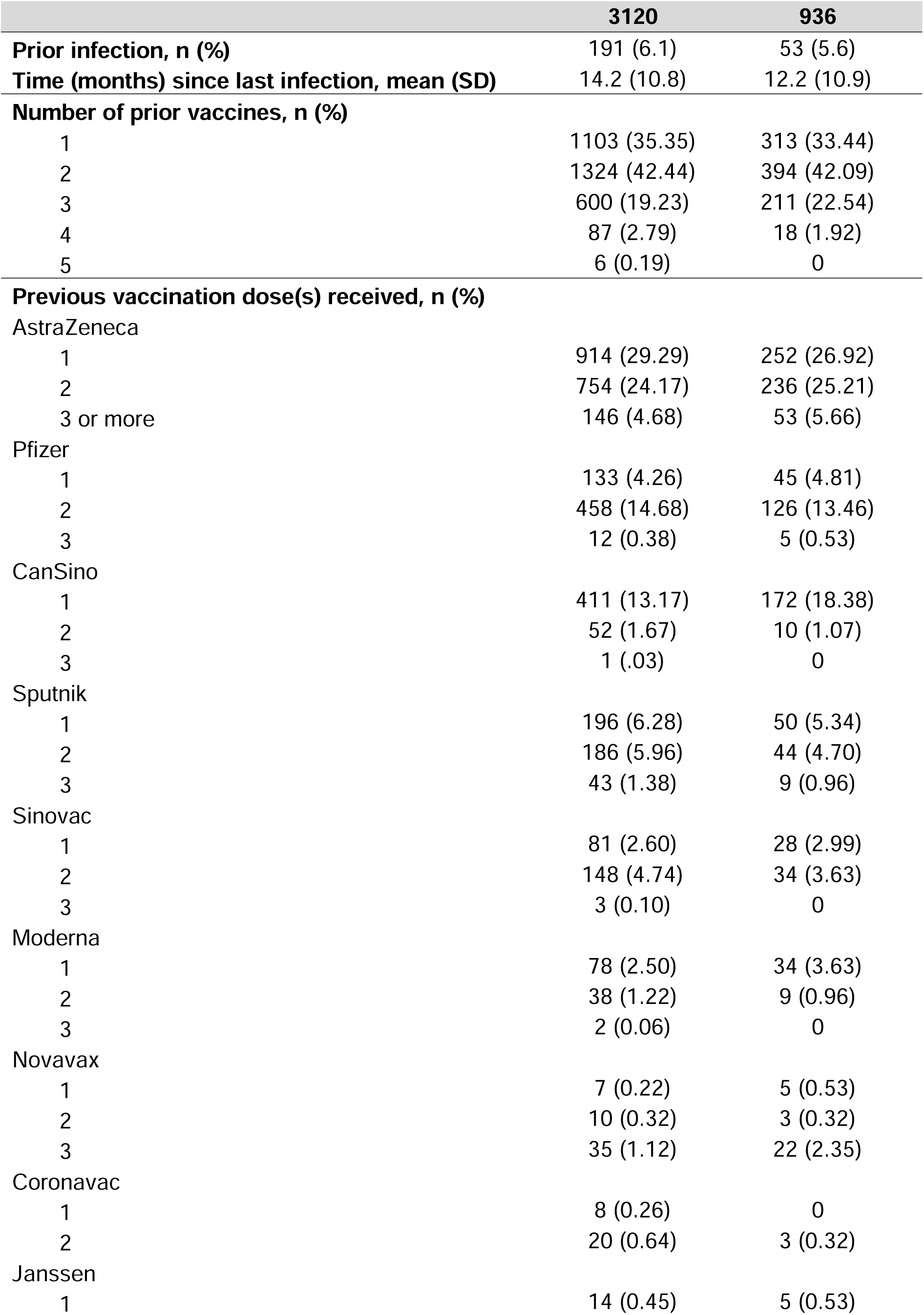
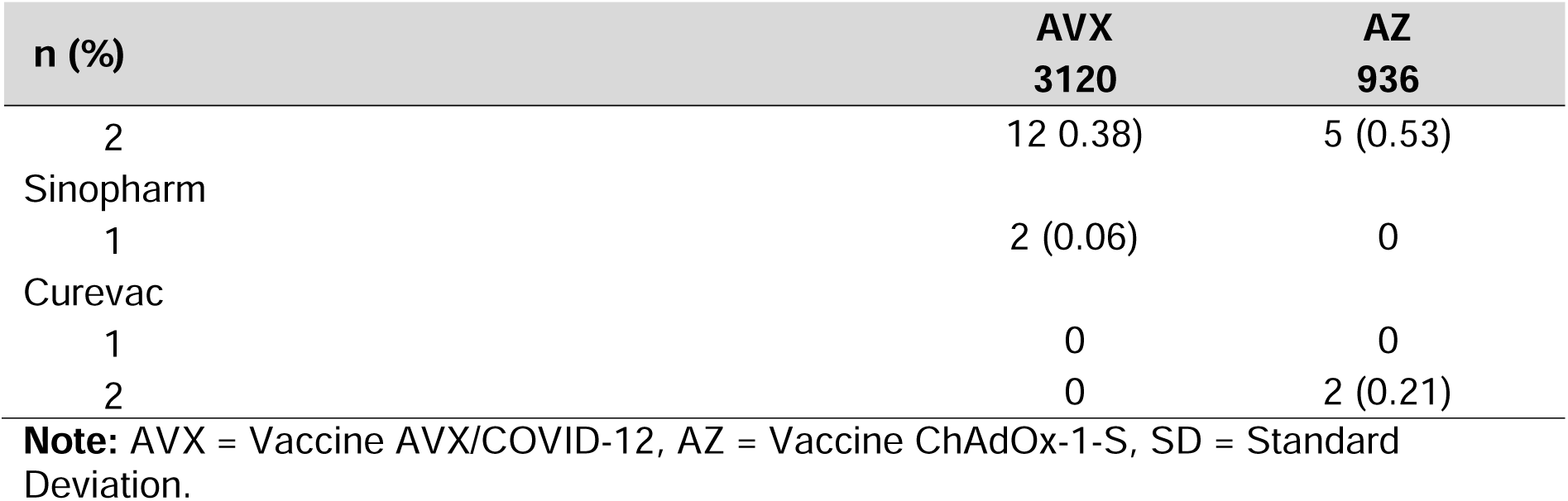
History of prior SARS-CoV-2 vaccination and/or infection of the phase II/III participants at baseline.

|  | 3120 | 936 |
| --- | --- | --- |
| **Prior infection, n (%)** | 191 (6.1) | 53 (5.6) |
| **Time (months) since last infection, mean (SD)** | 14.2 (10.8) | 12.2 (10.9) |
| **Number of prior vaccines, n (%)** |  |  |
| 1 | 1103 (35.35) | 313 (33.44) |
| 2 | 1324 (42.44) | 394 (42.09) |
| 3 | 600 (19.23) | 211 (22.54) |
| 4 | 87 (2.79) | 18 (1.92) |
| 5 | 6 (0.19) | 0 |
| **Previous vaccination dose(s) received, n (%)** |  |  |
| AstraZeneca |  |  |
| 1 | 914 (29.29) | 252 (26.92) |
| 2 | 754 (24.17) | 236 (25.21) |
| 3 or more | 146 (4.68) | 53 (5.66) |
| Pfizer |  |  |
| 1 | 133 (4.26) | 45 (4.81) |
| 2 | 458 (14.68) | 126 (13.46) |
| 3 | 12 (0.38) | 5 (0.53) |
| CanSino |  |  |
| 1 | 411 (13.17) | 172 (18.38) |
| 2 | 52 (1.67) | 10 (1.07) |
| 3 | 1 (.03) | 0 |
| Sputnik |  |  |
| 1 | 196 (6.28) | 50 (5.34) |
| 2 | 186 (5.96) | 44 (4.70) |
| 3 | 43 (1.38) | 9 (0.96) |
| Sinovac |  |  |
| 1 | 81 (2.60) | 28 (2.99) |
| 2 | 148 (4.74) | 34 (3.63) |
| 3 | 3 (0.10) | 0 |
| Moderna |  |  |
| 1 | 78 (2.50) | 34 (3.63) |
| 2 | 38 (1.22) | 9 (0.96) |
| 3 | 2 (0.06) | 0 |
| Novavax |  |  |
| 1 | 7 (0.22) | 5 (0.53) |
| 2 | 10 (0.32) | 3 (0.32) |
| 3 | 35 (1.12) | 22 (2.35) |
| Coronavac |  |  |
| 1 | 8 (0.26) | 0 |
| 2 | 20 (0.64) | 3 (0.32) |
| Janssen |  |  |
| 1 | 14 (0.45) | 5 (0.53) |

| **n (%)** | **AVX**<br>**3120** | **AZ**<br>**936** |
| --- | --- | --- |
| 2 | 12 (0.38) | 5 (0.53) |
| Sinopharm |  |  |
| 1 | 2 (0.06) | 0 |
| Curevac |  |  |
| 1 | 0 | 0 |
| 2 | 0 | 2 (0.21) |

**Note:** AVX = Vaccine AVX/COVID-12, AZ = Vaccine ChAdOx-1-S, SD = Standard Deviation.

### Safety after boosting with AVX/COVID-12

Out of the total population evaluated for safety (n=4,056 subjects), 2,111 vaccinated subjects reported adverse events during the study. Among them, 51.37% of the subjects in the AVX/COVID-12 group (n=1,603) and 54.27% in the AZ/ChAdOx-1-S group (n=508) experienced adverse events, showing no statistically significant differences among the groups (Figure 2A). When categorizing events by severity (mild, moderate, or severe), it was observed that for mild events, a higher proportion of subjects were affected in the AZ/ChAdOx-1-S group compared to the AVX/COVID-12 vaccine group (p=0.02). Conversely, for moderate adverse events, the proportion was higher in individuals who received the AVX/COVID-12 vaccine (p=0.01). Finally, for severe adverse events, no statistically significant differences were observed between the two groups (Figure 2A).

**Figure 2.**
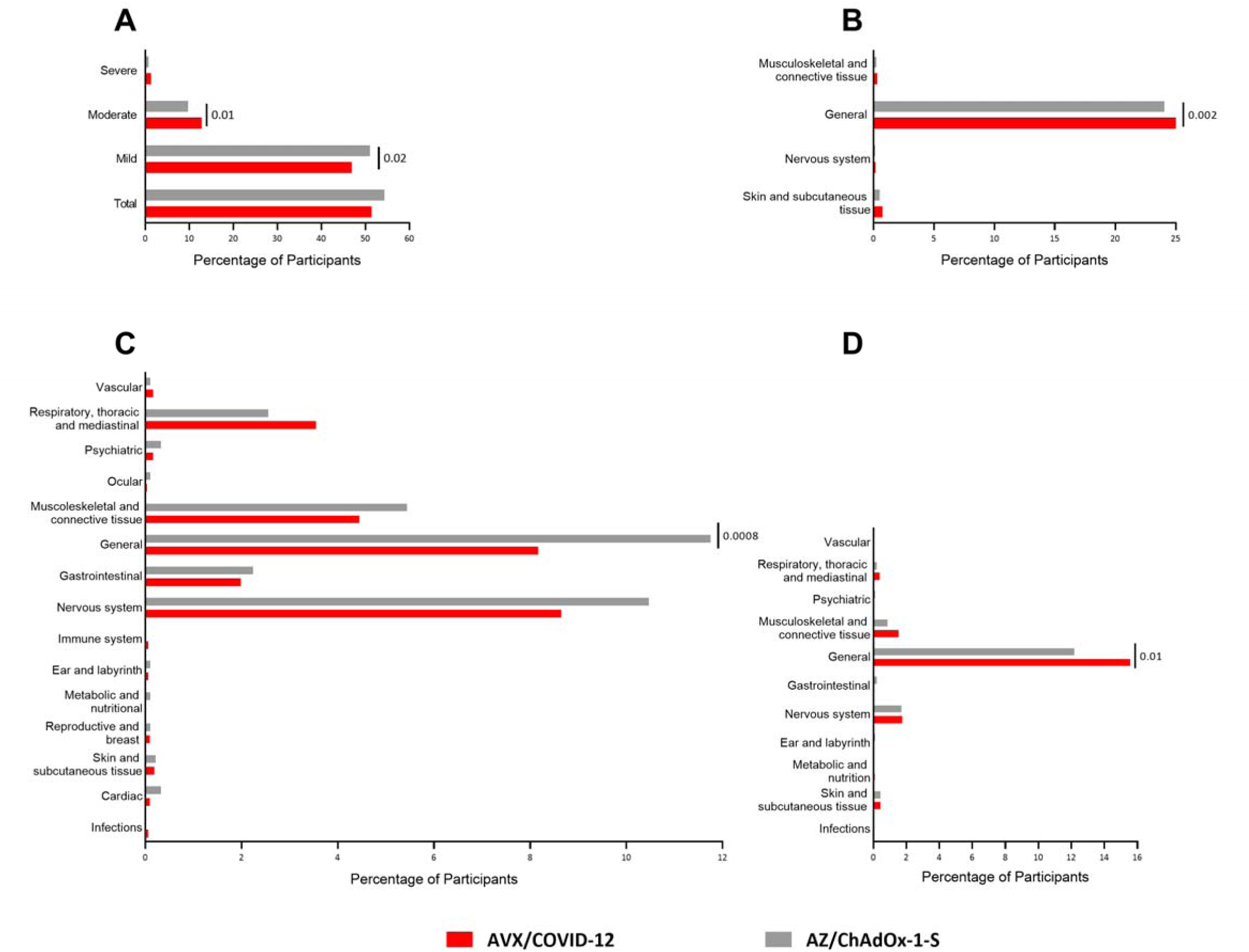
Local and systemic adverse events 7 days after boosting with AVX/COVID-12 or ChAdOx-1-S. Compilation of clinical data and adverse events in participants following boosting with AVX/COVID-12 or ChAdOx-1-S vaccines. A) Percentage of participants affected by adverse events classified by severity; B) participants affected by local adverse events of special interest; C) participants affected by systemic adverse events of special interest, and D) participants affected by adverse events with a confirmed relation to the vaccine according to the investigator’s criteria. The analyzed disorders are listed in suplementary tables 1 and 2. p-values were calculated through a comparison of proportions using the Z-test. Statistically significant differences were observed with p-values: p≤0.05.

In the context of local adverse events of special interest, 800 volunteers (25.64%) were documented in the AVX/COVID-12 vaccinated group, while 193 volunteers (20.61%) were recorded in the AZ/ChAdOx-1-S vaccinated group. This indicates a statistically significant difference (p=0.002), with a higher incidence observed in the AVX/COVID-12 group compared to the AZ/ChAdOx-1-S group. Regarding musculoskeletal and connective tissue disorders, nervous system disorders, and skin and subcutaneous tissue disorders, there were no statistically significant differences observed between the two groups (Figure 2B). The analyzed disorders include musculoskeletal and connective tissue issues (arthralgia, myalgia, back pain, pain in a limb, and limb discomfort), local general disorders (heat, pain, edema, redness, hematoma, swelling, induration, inflammation, nerve injury, itching at the injection site) nervous system disorders (paresthesia) and skin and subcutaneous tissue disorders (erythema, papule, and itching) (Table 1S).

For systemic AEs of special interest within 7 days post-vaccination, statistically significant differences were observed only in cases of mild severity. Specifically, 496 cases (15.89%) were reported in volunteers from the AVX/COVID-12 group, compared to 183 cases (19.55%) from the AZ/ChAdOx-1-S group (p=0.008) (Table S2). Importantly, there was a statistically significant difference in general disorders and administration site alterations in favor of the AVX/COVID-12 group (p=0.0008) (Figure 2C). The analyzed disorders include vascular (flushing, hypertensive crisis, and hypotension), respiratory, thoracic, and mediastinal (asthma, nasal congestion, dyspnea, pharyngolaryngeal pain, hiccups, allergic rhinitis, rhinorrhea, cough, and productive cough), and psychiatric (mood changes, confusional state, insomnia, and irritability), ocular (allergic conjunctivitis and photopsia), musculoskeletal and connective tissue (arthralgia, back pain, myalgia, ligament sprain, muscle spasms, and discomfort in limbs), systemic general disorders, and administration site alterations (asthenia, chest pain, influenza-like illness, shivering, fatigue, peripheral coldness, hyperthermia, inflammation, discomfort, general discomfort, chest tightness, pyrexia, decreased thirst, and feeling cold), gastrointestinal (diarrhea, abdominal distension, abdominal pain, upper abdominal pain, constipation, nausea, odynophagia, vomiting, and aphthous ulcer), and nervous system (headache, dizziness, migraine, paresthesia, drowsiness, and tremor), immune system (hypersensitivity), ear and labyrinth (vertigo), metabolism and nutrition (decreased appetite), reproductive and breast (polymenorrhea, heavy menstrual bleeding, and menstrual disorders), skin and subcutaneous tissue (rash, excessive sweating, skin lesions, and hives), cardiac (chest pain, palpitations, and tachycardia), and infections (oral herpes) (Table 2S).

Regarding participants who experienced AEs potentially associated with the vaccine, a total of 539 volunteers (17.27%) from AVX/COVID-12 and 122 from AZ/ChAdOx-1-S (13.03%) were reported. Significant differences (p=0.01) were observed in general disorders and administration site alterations (asthenia, pain at the injection site, edema at the injection site, erythema at the injection site, chill, fatigue, hematoma at the injection site, swelling, induration at the injection site, inflammation at the injection site, nerve injury at the injection site, discomfort, general discomfort, fever, and decreased thirst), predominantly occurring in volunteers vaccinated with AVX/COVID-12 (Figure 2D). No statistical differences were observed in the other analyzed disorders (Figure 2D).

### AVX/COVID-12 booster-induced neutralizing antibody titers against Wuhan-1 strain and Omicron SARS-CoV-2 variants of concern

The evaluation of neutralizing antibody titers in sera from AVX/COVID-12 boosted volunteers against the spike protein was conducted using the pseudovirus neutralization assay at 0, 14, 90, and 180 days post-immunization. Following the booster with AVX/COVID-12, anti-SARS-CoV-2 antibody titers against the ancestral Wuhan-1 strain or Omicron (BA.2 and BA.5) variants of concern were induced. Specifically, titers against the ancestral Wuhan-1 variant significantly increased at day 14 and slightly declined at day 90 and 180 post-immunization. In contrast, for the BA.2 and BA.5 variants, significantly higher neutralizing titers were observed at days 14, 90, and 180 after boosting (Figure 3A).

**Figure 3.**
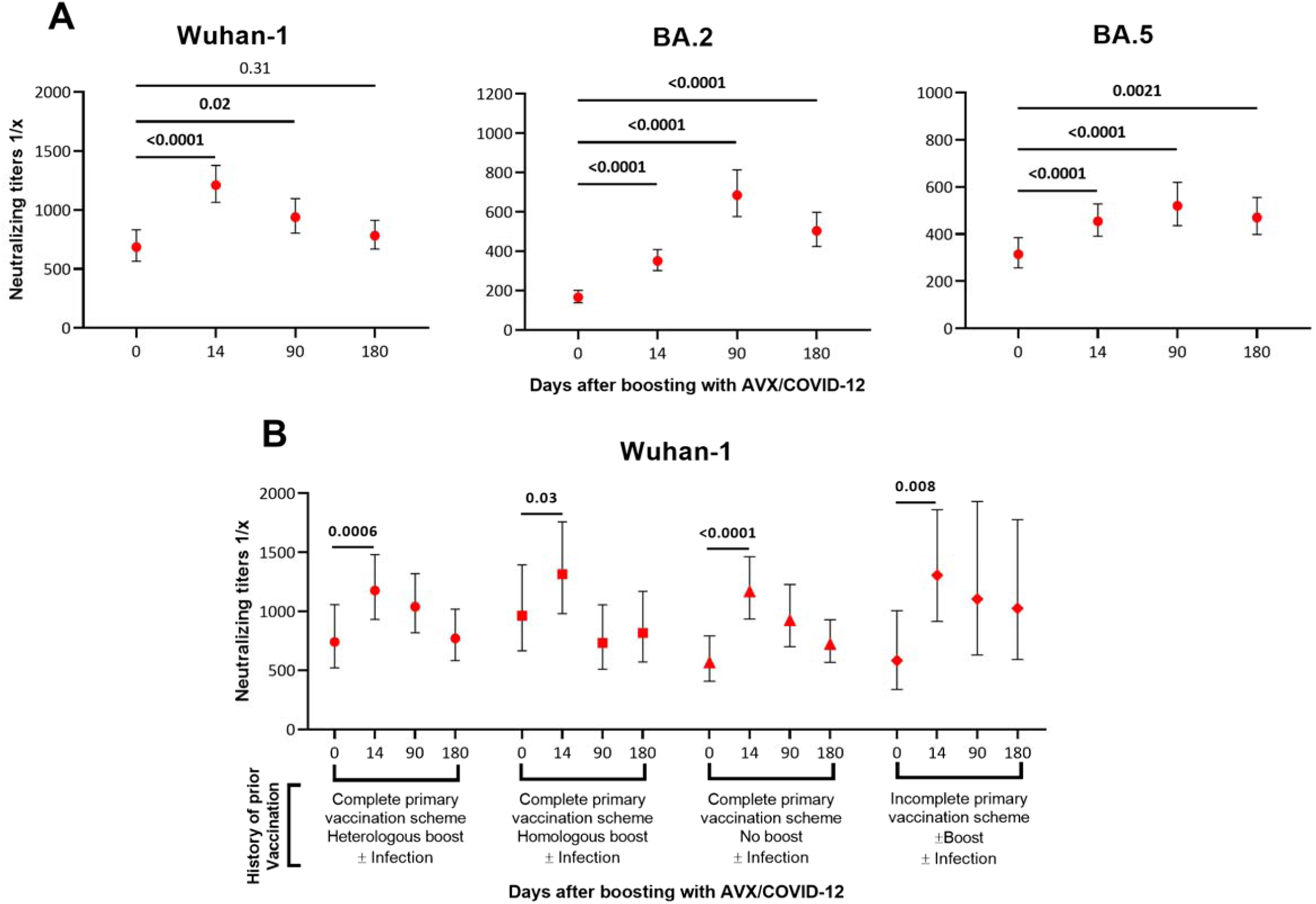
AVX/COVID-12 boosting induced neutralizing antibody titers against ancestral Wuhan-1 and Omicron SARS-CoV-2 variants of concern. Assessment of neutralizing antibody titers in sera from AVX/COVID-12 boosted volunteers against the S protein using the pseudovirus neutralization assay. A) Anti-SARS-CoV-2 antibody titers against Wuhan-1 or Omicron (BA.2 and BA.5) variants of concern. B) Specific neutralizing antibody titers against Wuhan-1 strain in participants with different history of immunization and infection induced by AVX/COVID-12 vaccine boost. Homologous boost refers to the use of the same vaccine platform used for the primary series vaccination; heterologous boost refers to the use a different vaccine platform used for the primary series vaccination. ± Infection groups includes infected and non-infected participants. The limit of detection is 60, although it is not specified in the graph. Dots represent the geometric mean, while bars indicate the 95% confidence intervals for the error. The p-values from the statistical analysis are presented in bold numbers, where p>0.05 is considered statistically significant. A paired t-test was employed to compare neutralizing titers.

The pattern of neutralizing titers remained consistent for BA.2 and BA.5 variants, depicting an increase at days 14 and 90, followed by a decline at day 180 post-boosting (Figure 3A). Notably, neutralizing antibody titers against the BA.2 and BA.5 variants were 1 to 3 times lower than those observed against the ancestral Wuhan-1 strain. Despite the decrease in titers at day 180, it is important to highlight that these titers against the BA.2 and BA.5 variants remained significantly higher compared to baseline levels before boosting with the AVX/COVID-12 vaccine.

As part of exploratory objectives, the magnitude of the increase in neutralizing titers at 0, 14, 90, and 180 days, following the administration of the AVX/COVID-12 vaccine in a single IM booster dose regimen, was analyzed. This analysis was stratified based on the immunization/infection history at the time of recruitment (Figure 3B). The data consistently demonstrate significantly increased titers at day 14 across all subgroups. However, following this time point, titers tended to decrease in all four subgroups. Based on this analysis, no differences were noted among the subgroups following the administration of the AVX/COVID-12 vaccine booster (Figure 3B). Here, “homologous boost” indicates using the same vaccine platform as the primary series, whereas “heterologous boost” refers to employing a different vaccine platform for the primary series. Analysis of the groups was conducted irrespective of their prior infection status (Figure 3B)

The neutralizing antibody titers following AVX/COVID-12 and AZ/ChAdOx-1-S boosting were evaluated across participants with comorbidities and different age groups. The antibody titers at day 0 and day 14 indicated that both vaccines induced similar enhancements in neutralizing antibody titers among participants with comorbidities and those aged > 65 and < 65 years (Figure 2S).

### Phase III immunobridging

#### Similar neutralizing antibody responses after booster doses of AVX/COVID-12 or AZ/ChAdOx-1-S vaccines

The non-inferiority of AVX/COVID-12, in comparison to the AZ/ChAdOx-1-S vaccine, was evaluated in 705 volunteers who received the AVX/COVID-12 booster and 712 who received the AZ/ChAdOx-1-S booster. Both groups were comparable, showing no statistical differences in age, gender, or race. However, there were statistically significant differences observed in individuals with obesity and normal body mass index (BMI), as well as those with 0 or 1 comorbidities. Nevertheless, no differences were noted among underweight and overweight volunteers or individuals with 1, 2, and 3 or more comorbidities (Table 3). Additionally, there were no statistically significant differences in volunteers based on their prior infection or vaccination history (Table 4).

**Table 3.**
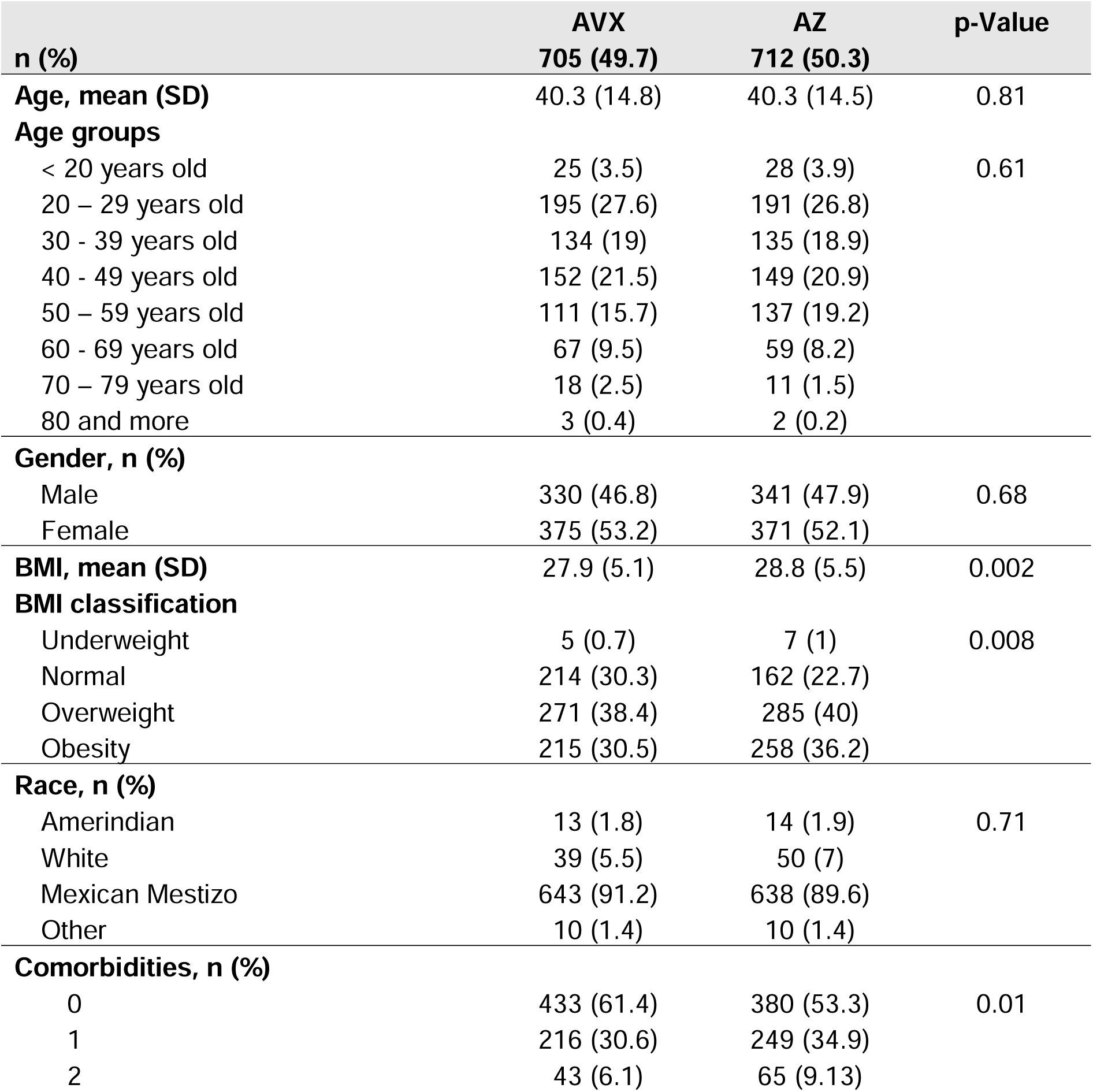
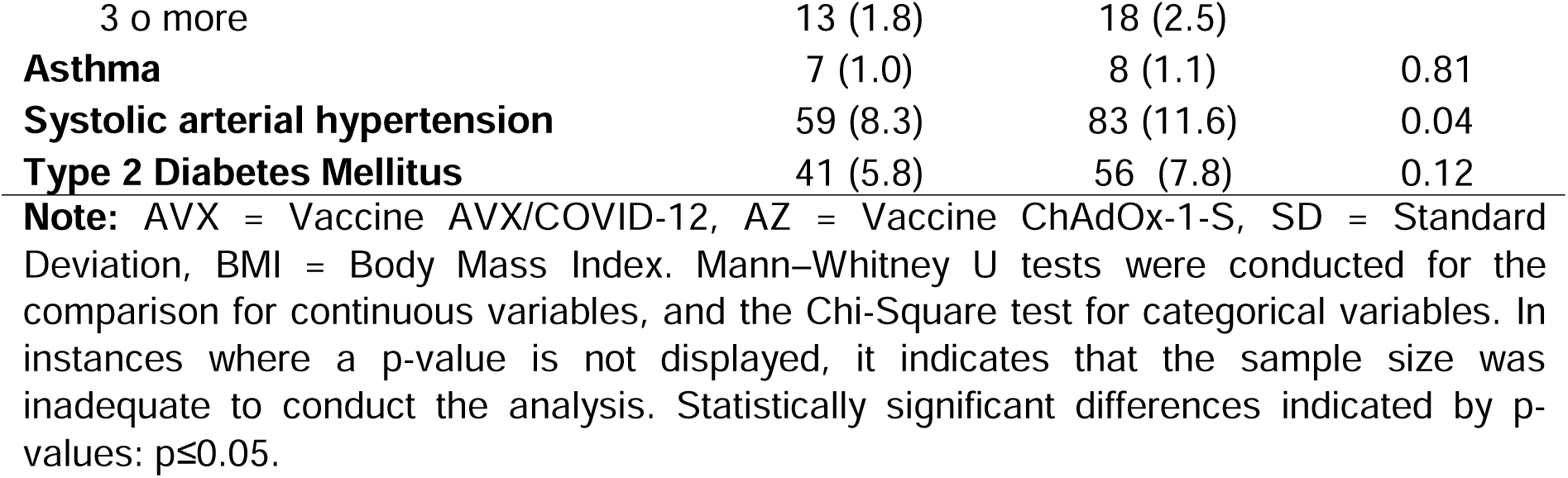
Demographic characteristics of phase III immunobridging.

| **n (%)** | **AVX**<br>**705 (49.7)** | **AZ**<br>**712 (50.3)** | **p-Value** |
| --- | --- | --- | --- |
| **Age, mean (SD)** | 40.3 (14.8) | 40.3 (14.5) | 0.81 |
| **Age groups** |  |  |  |
| < 20 years old | 25 (3.5) | 28 (3.9) | 0.61 |
| 20 – 29 years old | 195 (27.6) | 191 (26.8) |  |
| 30 - 39 years old | 134 (19) | 135 (18.9) |  |
| 40 - 49 years old | 152 (21.5) | 149 (20.9) |  |
| 50 – 59 years old | 111 (15.7) | 137 (19.2) |  |
| 60 - 69 years old | 67 (9.5) | 59 (8.2) |  |
| 70 – 79 years old | 18 (2.5) | 11 (1.5) |  |
| 80 and more | 3 (0.4) | 2 (0.2) |  |
| **Gender, n (%)** |  |  |  |
| Male | 330 (46.8) | 341 (47.9) | 0.68 |
| Female | 375 (53.2) | 371 (52.1) |  |
| **BMI, mean (SD)** | 27.9 (5.1) | 28.8 (5.5) | 0.002 |
| **BMI classification** |  |  |  |
| Underweight | 5 (0.7) | 7 (1) | 0.008 |
| Normal | 214 (30.3) | 162 (22.7) |  |
| Overweight | 271 (38.4) | 285 (40) |  |
| Obesity | 215 (30.5) | 258 (36.2) |  |
| **Race, n (%)** |  |  |  |
| Amerindian | 13 (1.8) | 14 (1.9) | 0.71 |
| White | 39 (5.5) | 50 (7) |  |
| Mexican Mestizo | 643 (91.2) | 638 (89.6) |  |
| Other | 10 (1.4) | 10 (1.4) |  |
| **Comorbidities, n (%)** |  |  |  |
| 0 | 433 (61.4) | 380 (53.3) | 0.01 |
| 1 | 216 (30.6) | 249 (34.9) |  |
| 2 | 43 (6.1) | 65 (9.13) |  |
| 3 or more | 13 (1.8) | 18 (2.5) |  |
| **Asthma** | 7 (1.0) | 8 (1.1) | 0.81 |
| **Systolic arterial hypertension** | 59 (8.3) | 83 (11.6) | 0.04 |
| **Type 2 Diabetes Mellitus** | 41 (5.8) | 56 (7.8) | 0.12 |

**Note:** AVX = Vaccine AVX/COVID-12, AZ = Vaccine ChAdOx-1-S, SD = Standard Deviation, BMI = Body Mass Index. Mann–Whitney U tests were conducted for the comparison for continuous variables, and the Chi-Square test for categorical variables. In instances where a p-value is not displayed, it indicates that the sample size was inadequate to conduct the analysis. Statistically significant differences indicated by p-values: $p \leq 0.05$ .

**Table 4.**
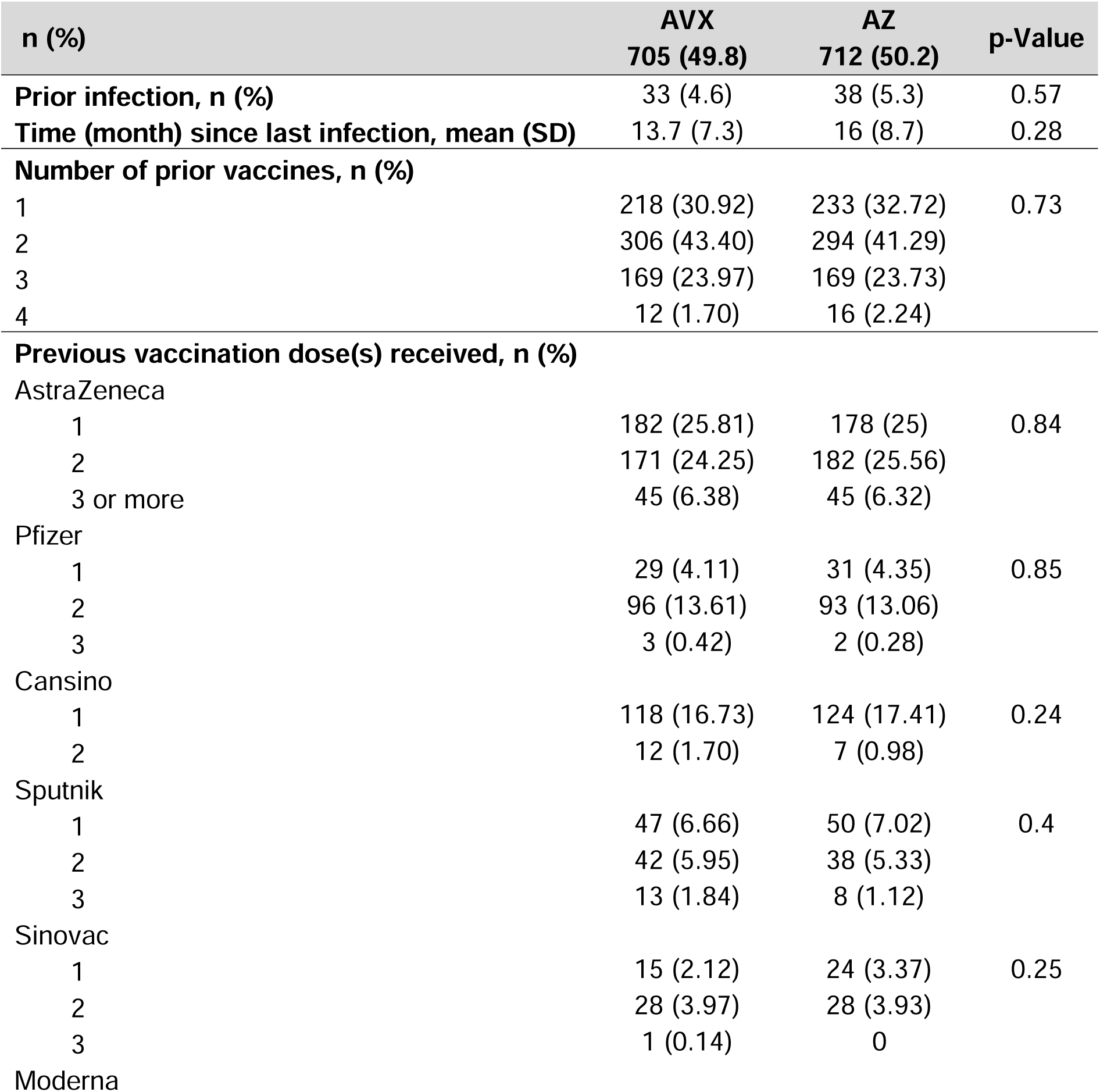
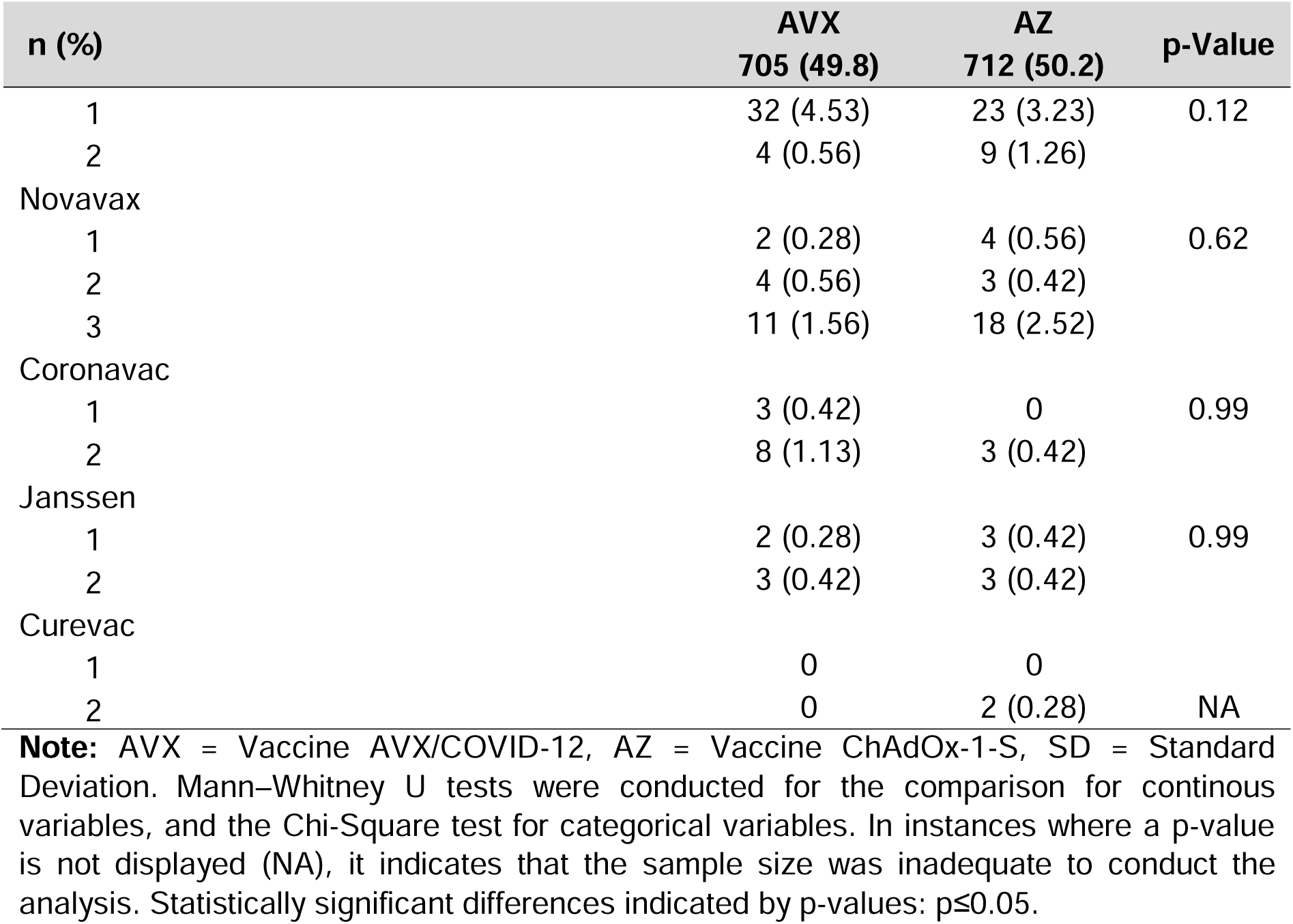
History of prior SARS-CoV-2 vaccination and/or infection of participants of the phase III immunobridging study at baseline.

The assessment of the non-inferiority of the AVX/COVID-12 vaccine, compared to the AZ/ChAdOx-1-S vaccine, was conducted by calculating the confidence interval for the ratio of geometric means of neutralizing titers and the confidence interval for the difference in the proportion of subjects with seroconversion, following the WHO guidelines for non-inferiority vaccine studies (19).

The analysis of the anti-Wuhan-1 strain neutralizing titers revealed well-balanced baseline values for the overall subject sample (p=0.23) (Table 5). The geometric mean ratio between those immunized with the AVX/COVID-12 vaccine and those with the AZ/ChAdOx-1-S vaccine at 14 days is 0.96, with a confidence interval of 0.85-1.06 (Table 5), demonstrating that AVX/COVID-12 is non-inferior to AZ/ChAdOx-1-S (Figure 4A). This result aligns with the non-inferiority criterion recommended by the WHO, indicating a lower limit of the confidence interval greater than or equal to 0.67 (21).

**Figure 4.**
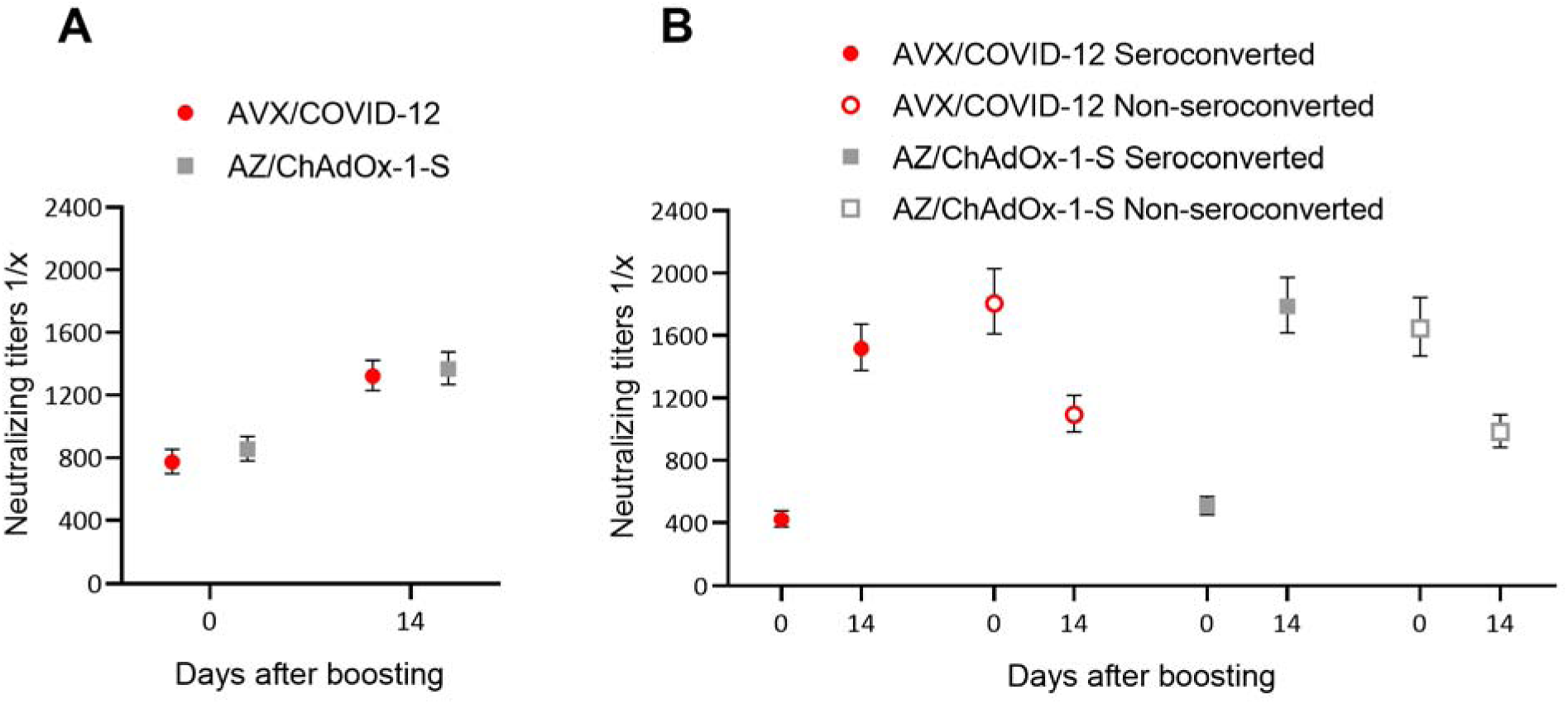
Similar neutralizing antibody responses were observed in participants who received booster doses of AVX/COVID-12 or AZ/ChAdOx-1-S. Analysis of neutralizing antibody titers against Wuhan-1 strain in participants boosted with AVX/COVID-12 (red circles) or AZ/ChAdOX-1-S (gray squares): A) Antibody titers in all boosted subjects on days 0 and 14 and B) comparison between participants who seroconverted (closed symbols) and subjects who did not seroconverted (open symbols). No statistically significant differences were observed, with p>0.05 considered as the threshold for statistical significance.

**Table 5.**
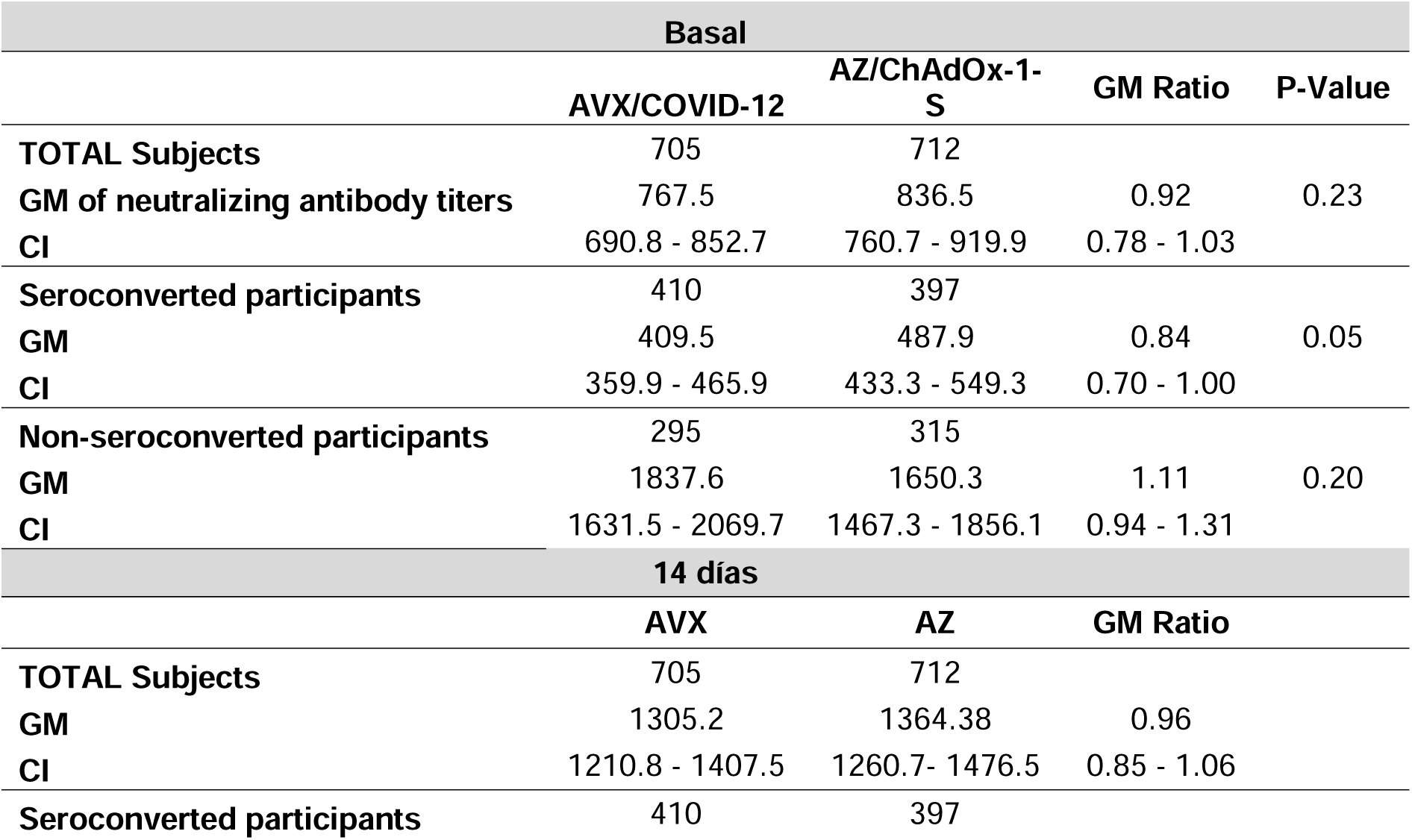
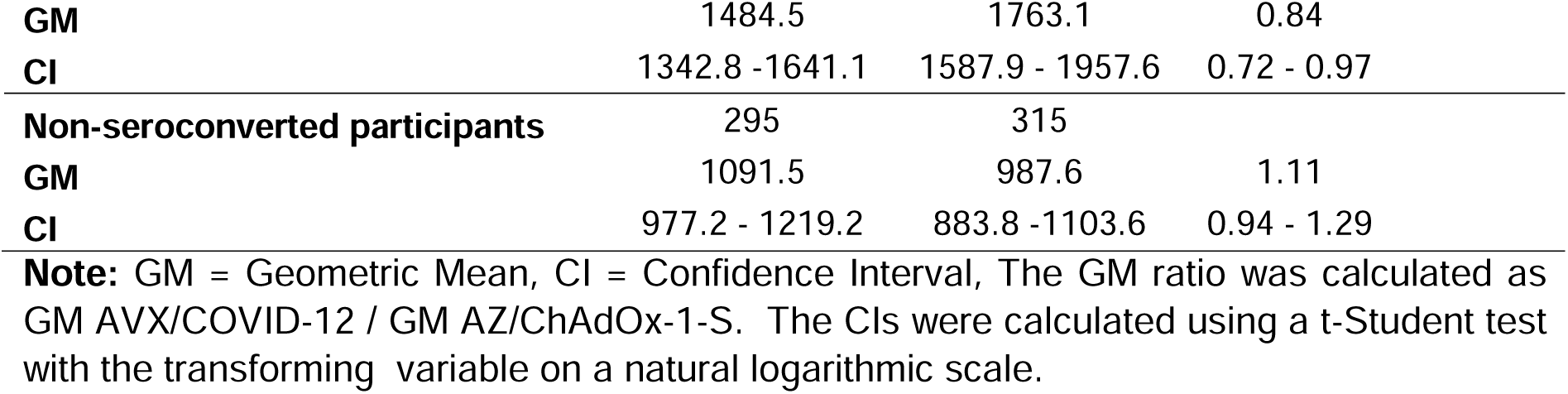
AVX/COVID-12 vaccine meets WHO’s non-inferiority criteria in comparison to AZ/ChAdOx-1-S neutralizing antibody titers at 14 days post-boosting.

| Basal |  |  |  |  |
| --- | --- | --- | --- | --- |
|  | AVX/COVID-12 | AZ/ChAdOx-1-S | GM Ratio | P-Value |
| TOTAL Subjects | 705 | 712 |  |  |
| GM of neutralizing antibody titers | 767.5 | 836.5 | 0.92 | 0.23 |
| CI | 690.8 - 852.7 | 760.7 - 919.9 | 0.78 - 1.03 |  |
| Seroconverted participants | 410 | 397 |  |  |
| GM | 409.5 | 487.9 | 0.84 | 0.05 |
| CI | 359.9 - 465.9 | 433.3 - 549.3 | 0.70 - 1.00 |  |
| Non-seroconverted participants | 295 | 315 |  |  |
| GM | 1837.6 | 1650.3 | 1.11 | 0.20 |
| CI | 1631.5 - 2069.7 | 1467.3 - 1856.1 | 0.94 - 1.31 |  |
| 14 días |  |  |  |  |
|  | AVX | AZ | GM Ratio |  |
| TOTAL Subjects | 705 | 712 |  |  |
| GM | 1305.2 | 1364.38 | 0.96 |  |
| CI | 1210.8 - 1407.5 | 1260.7- 1476.5 | 0.85 - 1.06 |  |
| Seroconverted participants | 410 | 397 |  |  |

|  |  |  |  |
| --- | --- | --- | --- |
| **GM** | 1484.5 | 1763.1 | 0.84 |
| **CI** | 1342.8 -1641.1 | 1587.9 - 1957.6 | 0.72 - 0.97 |
| **Non-seroconverted participants** | 295 | 315 |  |
| **GM** | 1091.5 | 987.6 | 1.11 |
| **CI** | 977.2 - 1219.2 | 883.8 -1103.6 | 0.94 - 1.29 |

**Note:** GM = Geometric Mean, CI = Confidence Interval, The GM ratio was calculated as GM AVX/COVID-12 / GM AZ/ChAdOx-1-S. The CIs were calculated using a t-Student test with the transforming variable on a natural logarithmic scale.

Upon further analysis of the neutralizing response, when stratifying boosted volunteers into seroconverted and non-seroconverted groups, it was found that 58.15% of subjects in the AVX/COVID-12 vaccinated group seroconverted, while in the AZ/ChAdOx-1-S vaccinated group, the proportion was 55.75%. The AVX/COVID-12 vaccinated group maintains a 2.4% advantage over the AZ/ChAdOx-1-S vaccinated group, with a confidence interval (CI) of −2.7 to 7.5. This outcome is consistent with the protocol non-inferiority criterion, indicating that the difference in both percentages should not exceed -10 (21). Notably, individuals in both vaccine groups who experienced seroconversion exhibited an approximately fourfold increase in their neutralizing titers 14 days after immunization. Conversely, the group that did not undergo seroconversion showed a decline in neutralizing titers at day 14 after boost for both vaccines (Figure 4B). This group was characterized by individuals with high titers of neutralzing antibodies prior to the vaccination boost, and these titers still remained high after the observed decline and day 14 after the boost (Figure 4B)

### T-cell responses among participants immunized with AVX/COVID-12 or AZ/ChAdOx-1-S vaccines

The production of IFN-γ by peripheral blood total, CD4+, and CD8+ T cells in response to stimulation with the subunit 1 of the spike protein of SARS-CoV-2 was assessed on days 0, 14, 90, and 180 in PBMCs from volunteers who received booster doses of AVX/COVID-12 (Figure 5A) or AZ/ChAdOx-1-S (Figure 5B), using flow cytometry.

**Figure 5:**
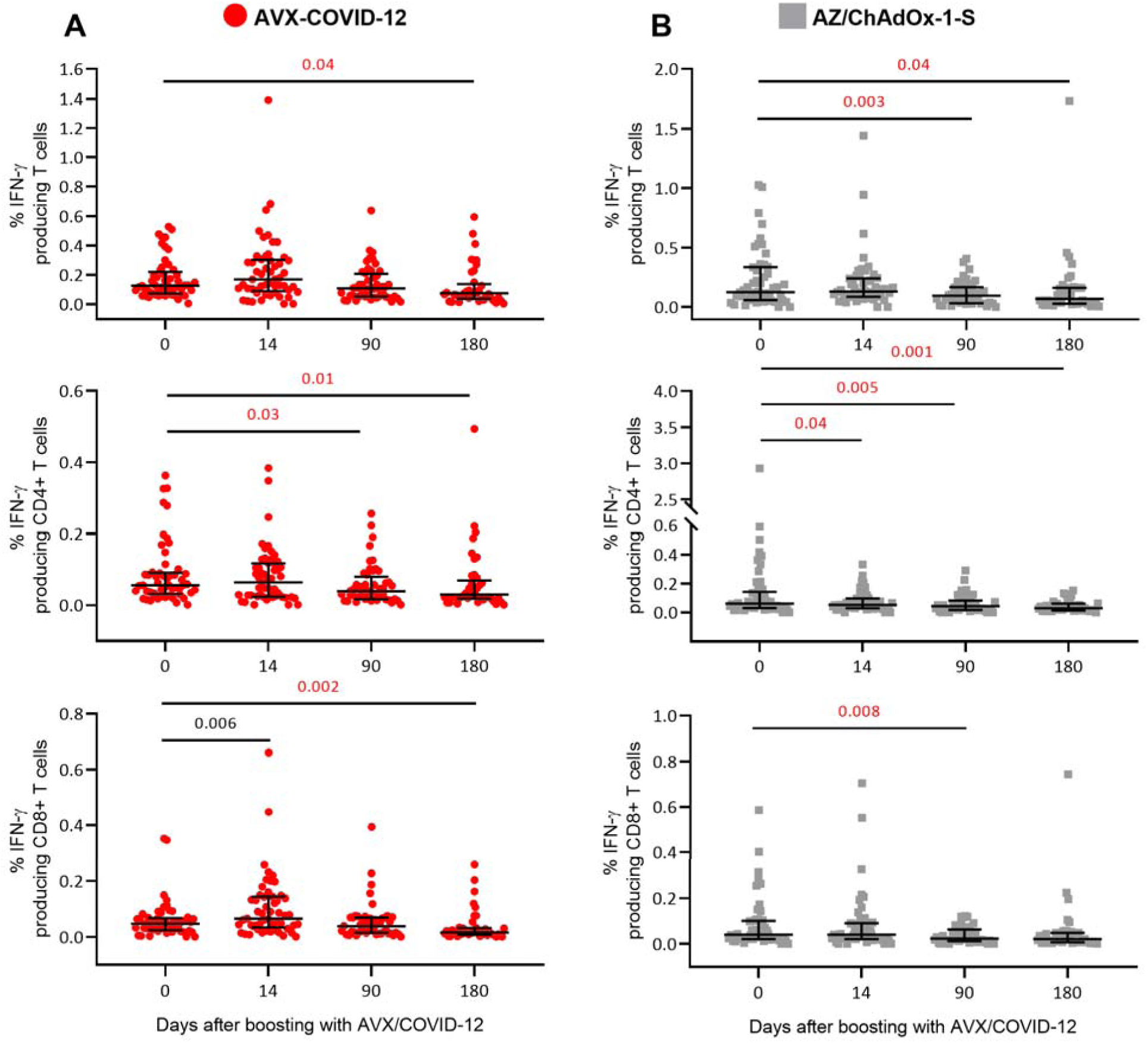
T cell responses in participants vaccinated with AVX/COVID-12 or AZ/ChAdOx-1-S vaccines. A) Percentage of IFN-γ- producing total, CD4+ and CD8+ T cells from peripheral blood mononuclear cells collected from participants who received booster doses of AVX/COVID-12 or B) AZ/ChAdOx-1-S vaccines. The cells were stimulated with the subunit 1 of spike protein, and the production of IFN-γ was assessed using flow cytometry. The limit of detection (LoD) of the % IFN-γ producing total T cells is 0.006, % IFN-γ producing CD4+ T cells is 0.006 and % IFN-γ producing CD8+ T cells is 0.006. The graph shows individual value. Bars shows geometric Mean, and the error shows the 95% confidence intervals. Statistically significant increases are depicted in black; this was observed only in IFN-γ-producing T cells from participants vaccinated with AVX/COVID-12. Statistically significant differences with decreases are shown in red. p-values: p≤0.05 for intragroup comparisons using the Wilcoxon Friedman signed-rank test.

In subjects vaccinated with AVX/COVID-12, a slight increase in total T cells and CD4+ T cells producing IFN-γ was observed on day 14; however, this rise did not reach statistical significance. Nevertheless, a notable increase in the percentage of IFN-γ-producing CD8+ T cells (p=0.006) was detected on day 14 compared to the baseline levels on day 0 (Figure 5A and Table 3S). Statistically significant decreases were observed in total IFN-γ-producing T cells only at day 180. In the case of the IFN-γ-producing CD4+ T cells, a decrease was noted at day 90 with no further decrease. For IFN-γ-producing CD8+ T cells, the reduction was observed only at day 180 (Figure 5A and Table 3S).

For volunteers immunized with AZ/ChAdOx-1-S, no significant increases were noted in total CD4+ and CD8+ IFN-γ-producing T cells (Figure 5B and Table 3S). Statistically significant decreases in IFN-γ-producing T cells were observed only at days 90 and 180. In the case of the IFN-γ-producing CD4+ T cells, the decrease occurred at day 14, with no further drop observed. Finally, a reduction in IFN-γ- producing CD8+ T cells was observed at day 90, with no subsequent change (Figure 5B and Table 3S). In Figure 5, statistically significant increases are depicted in black, and decreases are shown in red.

### Similar incidence of COVID-19 cases in volunteers boosted with AVX/COVID-12 or AZ/ChAdOx-1-S vaccines

As part of the secondary objectives for phase III, the occurrence of symptomatic COVID-19 cases was documented for both the AVX/COVID-12 and AZ/ChAdOx-1-S groups of boosted volunteers during the 14 days following administration. A total of 37 COVID-19 cases (5.5%) were recorded in the AVX/COVID-12 boosted group, and 42 cases were reported in the AZ/ChAdOx-1-S group (6.3%), resulting in an incidence rate per 1000 days of 0.29 and 0.32 for AVX/COVID-12 and AZ/ChAdOx-1-S, respectively (Table 6). Despite the trial not being designed for efficacy analysis, the log rank test indicated no significant difference in the cumulative incidence of COVID-19 cases between both groups (p=0.42) (Table 6). Additionally, a comparison of Nelson-Aalen cumulative hazard incidence curves for COVID-19 cases reported during the 180 days after boosting with AVX/COVID-12 or AZ/ChAdOx-1-S showed no statistically significant differences between volunteers vaccinated with AVX/COVID-12 or AZ/ChAdOx-1-S (Figure 6).

**Figure 6.**
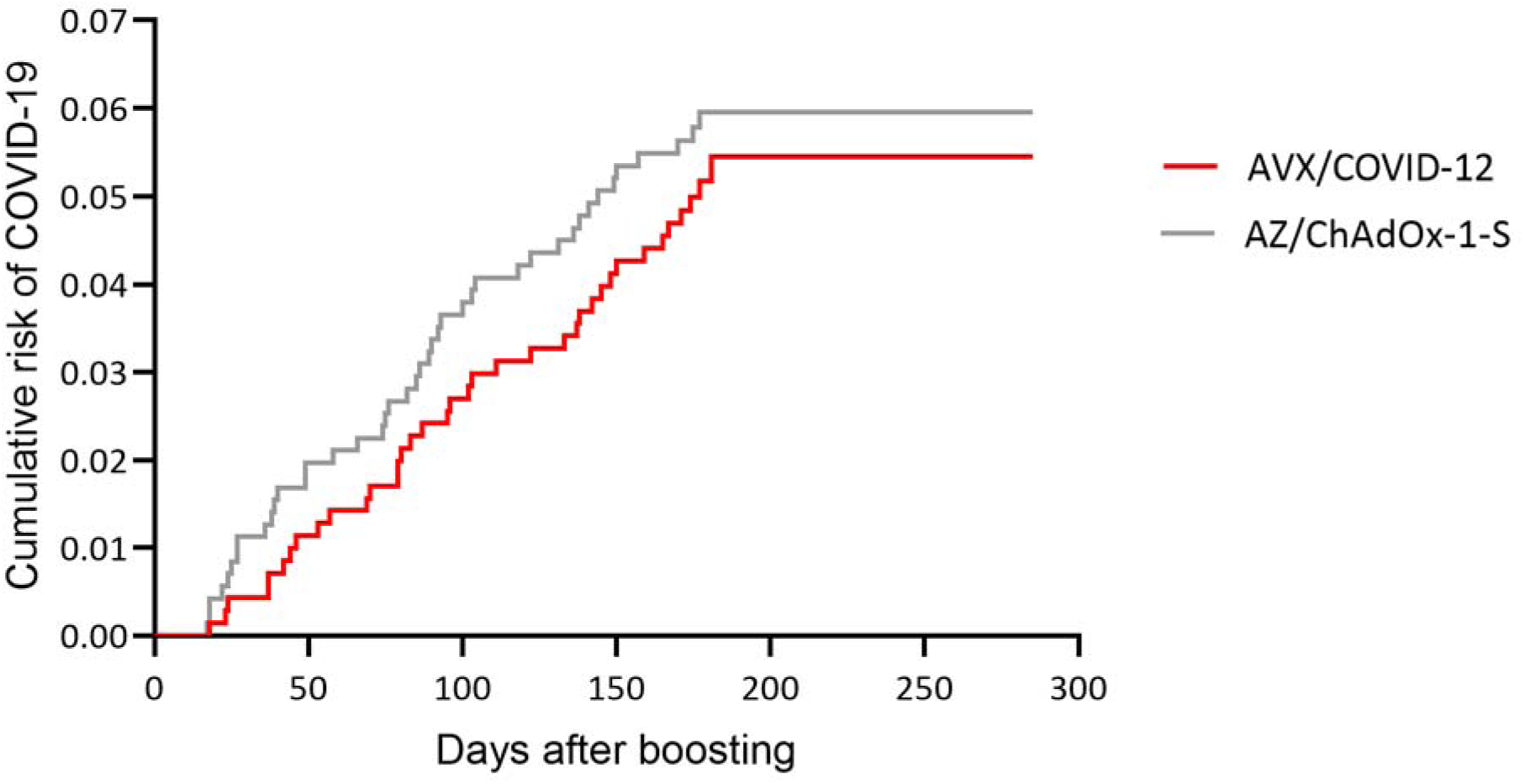
The incidence of COVID-19 cases showed no statistically significant difference between the participants vaccinated with AVX/COVID-12 or AZ/ChAdOx-1-S. Comparison of Nelson-Aalen cumulative hazard incidence curves for COVID-19 cases reported during the 180 days after boosting with AVX/COVID-12 or AZ/ChAdOx-1-S.

**Table 6.** Incidence of COVID-19 cases.

| **Subjects in the study** | **Total**<br>**1,417** | **AVX/COVID-12**<br>**705 (49.8)** | **AZ/ChAdOx-1-S**<br>**712 (50.2)** |
| --- | --- | --- | --- |
| Total follow-up time in days | 255,143 | 127,120 | 128,023 |
| Average time (SD) in days | 180 (31.6) | 180.3 (29.6) | 179.8 (33.5) |
| COVID-19 cases (%)<sup>1</sup> | 79 (5.5) | 37 (5.3) | 42 (6.3) |
| Incidence rate per 1000 days | 0.30 | 0.29 | 0.32 |

<sup>1</sup>Log Rank Test: p-value = 0.58

## Discussion

The naturally attenuated lentogenic NDV LaSota strain is an avian virus that exhibits host-range restriction, thereby preventing productive multicycle infection in non-avian species, including humans. As a live vaccine vector, it has been employed in the veterinary industry for over 70 years, demonstrating a commendable track record of safety and efficacy (22). NDV-vectored vaccines have undergone preclinical studies against various coronaviruses, revealing their safety and immunogenicity in natural poultry host and non-natural mammalian challenge models (23–25). The natural attenuation of NDV confers an advantage over other live viral vaccines used in humans, as it is unlikely to induce disease in humans or any wild or domestic avian species. Studies have indicated that the introduction of foreign genes into NDV genomes results in reduced pathogenicity in birds, rather than an increase (26). Consequently, the NDV-based AVX/COVID-12 does not pose any discernible environmental or agricultural risks (27). Of note, the inserted HexaPro spike protein is non-functional since its fusion machinery has been disabled by the introduction of the prolines and by removing the cleavage site between spike subunit 1 and spike subunit 2.

The safety of the AVX/COVID-12 vaccine, administered via IM or IN routes, was initially confirmed through preclinical tests conducted in pigs (10). This confirmation was validated in a phase I clinical trial involving volunteers with no prior history of infection or vaccination (28). Considering that a significant portion of the population had previously encountered infection or received vaccination at the time of the study, a subsequent phase II clinical trial was conducted to assess the safety and immunogenicity of the AVX/COVID-12 vaccine as a heterologous booster dose through both IM and IN routes. The trial demonstrated that the vaccine retained its safety and immunogenicity when employed as a booster dose (16). Reports from clinical trials involving COVID-19 NDV-based vaccines similar to AVX/COVID-12 also indicate that these vaccines are safe and well tolerated in humans (12–15).

In this study, we further analyzed the safety and immunogenicity of the AVX/COVID-12 vaccine as an IM booster in healthy adult volunteers who have gone more than four months since their last COVID-19 vaccination and/or SARS-CoV-2 infection. Additionally, a non-inferiority study was conducted using the AZ/ChAdOx-1-S vaccine as an active control. The safety, immunogenicity, and efficacy of the intranasal administration of the AVX/COVID-12 vaccine will be investigated in future studies.

The assessment of AEs and their potential association with vaccination was conducted in 4,056 subjects over a six-month period. Of these, 3,120 received a single IM dose of the AVX/COVID-12 vaccine, while 936 received the AZ/ChAdOx-1-S vaccine as an active control. While the use of a placebo during the phase II stages facilitates a clear definition of the immune response magnitude in subjects receiving the experimental vaccine, incorporating placebo controls in pharmaceutical development protocols for COVID-19 vaccines, at the time of this trial, presented an ethical conflict as vaccines with emergency authorization and national vaccination plans became available. Consequently, placebo controls were not employed in this study. Instead, AZ/ChAdOx-1-S as active control was utilized during phase II to prevent bias in subject distribution and serve as a comparator for assessing the vaccine’s safety. On the other hand, during the phase III stage, the active control was employed to accurately define biologically relevant non-inferiority and avoid a potential bio-creep effect (29).

The primary vaccine-associated AEs in the phase II-III study were predominantly mild and associated with the administration site. Few severe AEs were reported, no statistically significant differences were observed between the two groups, and they were not related to vaccination. However, regarding the statistically significant differences, it was found that general reactions and pain at the injection site were more frequent in the AVX/COVID-12 group than in the AZ/ChAdOx-1-S group. Nevertheless, systemic events were more frequent in the case of the AZ/ChAdOx-1-S group. It is important to emphasize that there were no statistical differences in adverse events between the entire population vaccinated with AZ/ChAdOx-1-S or AVX/COVID-12 (Figure 2A). Furthermore, the nature, incidence, and severity of adverse events recorded during this study do not indicate any alarming signals following the administration of the AVX/COVID-12 vaccine. The demonstrated safety of the AVX/COVID-12 vaccine aligns with the outcomes of earlier clinical phases (11,16).

The findings outlined in this report highlight the immunogenicity of the vaccine, evidenced by an increase in neutralizing antibody titers, not only against the ancestral Wuhan-1 strain but also against the Omicron BA.2 and BA.5 variants of concern (Figure 3). While the neutralizing antibody responses showed an initial increase at day 14 followed by a gradual decline, it is noteworthy that even after 6 months, antibody titers remained higher compared to pre-boosting levels (Figure 3). This persistence aligns with observations reported for other COVID-19 vaccines (30).

Furthermore, the analysis of the geometric mean ratio (GMT) of neutralizing antibodies demonstrated that the AVX/COVID-12 experimental vaccine (GMT ratio: 0.96, CI: 0.85-1.06) met the standard set by the WHO (GMT≥0.67) (Table 5) (21), confirming its non-inferiority when compared to the AZ/ChAdOx-1-S vaccine.

The efficacy of the AZ/ChAdOx-1-S vaccine had been previously established in clinical studies with placebo controls (31). Moreover, primary-series AZ/ChAdOx-1-S vaccination protects against COVID-19 hospitalization with enduring levels of vaccine effectiveness through ≥6 months, as indicated by consistent results in a meta-regression analysis. The analysis showed ≥80% protection against COVID-19 hospitalization through approximately 43 weeks post-second dose, with a certain decline (17,18). This vaccine was the most widely used in vaccination campaigns in Mexico and is the most utilized globally, distributed in 185 countries (2,17). Therefore, it serves as an excellent active control for the non-inferiority study of the AVX/COVID-12 vaccine.

Further data analysis indicates that a percentage of individuals in both groups did not seroconvert. Approximately 40% of those vaccinated with both AVX/COVID-12 and AZ/ChadOx-1-S did not seroconvert when measuring response against the ancestral Wuhan-1 strain, while it was 20% when considering those who did not seroconvert in response to Wuhan, BA.2, and BA.5 (triple negatives). In other words, 60% seroconverted when measuring the response against the Wuhan-1 strain, and around 80% seroconverted when considering the response measured against Wuhan-1 or the BA.2 or BA.5 Omicron strains. This trend was observed in both the AVX/COVID-12 and AZ/ChadOx-1-S vaccinated groups (Figure 4B).

The lack of seroconversion has been reported both in some infected patients (32) and vaccinated individuals (33), particularly in patients with prior infection, where seroconversion after receiving a vaccine booster is much lower or non-existent when neutralizing titers are high due to previous infections (33). In this study, 32% of the volunteers exhibited anti-N titers (data not shown), indicating prior infection (or vaccination with inactivated vaccines). Additionally, the study was conducted during a pandemic peak caused by the Omicron variant (Figure 1S), leading to around 40% of participants having titers of 1,024 and above at baseline (Figure 4B). The analysis, comparing baseline titers and the percentage of seroconversion, clearly shows that higher baseline titers are associated with lower seroconversion, aligning with existing literature (33).

In the case of volunteers who did not seroconvert in both the AVX/COVID-12 and AZ/ChadOx-1-S boosted groups, it was interestingly observed that the antibody titers were declining at day 14 (Figure 4B). Further analysis revealed that these volunteers had very high baseline titers. This phenomenon has been previously reported in similar cases (33,34), especially in volunteers with high baseline titers receiving homologous boosters. Additionally, studies in mice have shown that immunization schemes involving multiple doses lead to a reduced response of neutralizing antibodies, with a diminished T-cell response, decreased formation of germinal centers, and cellular response exhaustion (35). This information could contribute to explaining the observed response in the study patients. It is also crucial to consider that most of these volunteers had already developed robust hybrid immunity due to previous vaccinations and infections. Furthermore, the study was conducted during a peak of Omicron infections (Figure 1S), and undetected exposure to the virus could have contributed to the observed response. These data indicate that in individuals with high neutralizing antibody titers, the boosting does not improve the response; on the contrary, it could not prevent waning in antibody titers. This information is relevant for planning and deploying vaccination campaigns to maximize the benefits of vaccination.

Nevertheless, the analysis of the volunteerś neutralizing antibody responses confirmed that the AVX/COVID-12 vaccine meets the WHO non-inferiority standard compared to the AZ/ChAdOx-1-S vaccine. Individuals with baseline neutralization titers around 400, boosted either with AVX/COVID-12 or AZ/ChAdOx-1-S, exhibited a 3 to 4-fold increase in seroconversion for both groups (Figure 4A and 4B). These results underscore the effectiveness and importance of timely boosting in specific individuals. It is important to note that this trial included participants regardless of their initial anti-S IgG titers. The seroconversion observed in volunteers with initially low antibody titers aligns with phase II clinical trial findings, where participants had similarly low anti-S IgG titers at the beginning of the study (16). Notably, this phase II/III study demonstrates that boosting individuals with high neutralizing antibody titers does not result in a further increase; in fact, it might not stop antibody waning, though this decline does not reach initial levels as high antibody titers persist.

The primary focus of anti-COVID-19 boosting campaigns is on the elderly, immunocompromised, and individuals with comorbidities. Upon further examination of neutralizing antibody titers following AVX/COVID-12 or AZ/ChAdOx-1-S boosting, both vaccines exhibited comparable enhancements in neutralizing antibody titers among participants with comorbidities, as well as those aged > 65 and < 65 years (Figure 2S). This data supports the use of these vaccines as boosters in these populations. Further studies are required to evalaute the safety and immunogenicity of this vaccine in immunocompromised indviduals, pregnant women and kids.

With time, the combination of infection and vaccination generates hybrid immunity, which has been demonstrated to broaden and enhance the neutralizing antibody and T-cell responses, providing increased protection against infection, hospitalization, and death (36). This report illustrates the induction of a neutralizing response against the Omicron variant of concern (VOC) through boosting with AVX/COVID-12 or AZ/ChAdOx-1-S. Since both vaccines express the spike protein of the ancestral Wuhan-1 strain, the anti-Omicron neutralizing response may be attributed to extensively documented cross-reactivity. This phenomenon is particularly evident in the context of hybrid immunity, which has been shown to broaden and enhance the response of neutralizing antibodies, not only against the ancestral variant but also against new variants and other seasonal coronaviruses as well as SARS-CoV-1 and the Middle East respiratory syndrome coronavirus (MERS CoV) (33). Given the pandemic status when the trial was conducted, the induction of antibodies against variants other than Wuhan-1 is not surprising, considering the large number of T and B cell epitopes shared by the spike proteins from SARS-CoV-2 variants of concern (37).

Regarding the cellular response, an increase in the total CD4+ and CD8+ T cell responses was observed on day 14 after receiving the booster dose. However, the increase was statistically significant only for CD8+ cells producing IFN-γ. It is important to note that the data suggest that at the initiation of this study, the proportions of IFN-γ-producing T lymphocytes were relatively high in the participating subjects, as there is a trend indicating a significant decrease at days 90 and 180. This trend is also observed for subjects enrolled in the group that received the AZ/ChAdOx-1-S vaccine, suggesting that, at a population level, due to the active circulation of the virus, there were initially high baseline levels of T lymphocytes specific to the SARS-CoV-2 spike protein. While it was possible to detect significant increases in the specific cellular response to the virus, the high baseline levels prevented clearer observations of cellular response increases (Figure S1). Considering the inclusion criteria, which allowed the recruitment of individuals vaccinated or previously infected with SARS-CoV-2 for four months or more, it is anticipated that the cellular response would already be elevated (30). This makes it challenging to observe a distinct increase after the booster. Reports from 2022 estimated that at least 51% of the population had hybrid immunity, with 85% exhibiting some level of Omicron-induced antibody titers (38). Additional studies propose limited potential for further boosting, notably in individuals initially primed with infection followed by three mRNA vaccine doses, which maximally induce spike-specific T-cell responses (39). This report also illustrates that a previous history of SARS-CoV-2 infection can modify immune responses to the spike protein during Omicron infection in vaccinated populations. Nevertheless, Omicron infection in individuals who have received triple vaccinations generally heightens immune responses to SARS-CoV-2 in both the bloodstream and mucosal regions. This enhancement potentially fortifies long-lasting population immunity against SARS-CoV-2 (39). Other studies have indicated that vaccination with BNT162b2, based on the Wuhan-1 spike protein, and/or breakthrough infection by early Omicron subvariants elicits CD8+ T cell responses that recognize epitopes within the spike protein of newly emerging subvariants (40).

Collectively, these findings suggest that hybrid immunity acquired from both vaccination and past infections can prompt notably heightened and broader immune responses. This combined immunity shows the potential to offer protection against emerging variants of concern.

The evaluation of COVID-19 cases among recipients of AVX/COVID-12 or AZ/ChAdOx-1-S vaccines revealed no cases of severe disease, with only mild to moderate disease observed in both groups. An analysis conducted 14 days post-vaccination indicated a trend towards a lower occurrence of COVID-19 cases in individuals boosted with AVX/COVID-12 in comparison to the AZ/ChAdOx-1-S vaccine group, despite the trial not being designed for efficacy analysis.

The urgent need persists for the development of safe, effective, and cost-efficient vaccines against COVID-19, especially for implementing booster campaigns aimed at vulnerable groups like the elderly, immunocompromised individuals, and those with comorbidities. This also serves the crucial goal of limiting transmission among the wider population. The results presented here collectively support the utilization of the AVX/COVID-12 vaccine to address this pressing need.

## Conclusions

Our findings demonstrate that the AVX/COVID-12 vaccine is safe, well-tolerated, and highly immunogenic. It effectively stimulated the production of neutralizing antibodies against the spike protein from both the ancestral Wuhan-1 strain and the Omicron variants BA.2 and BA.5. Additionally, at day 14 post-immunization, AVX/COVID-12 uniquely induced IFN-γ-producing CD8+ T cells. Furthermore, a discernible trend indicated a potentially lower incidence of COVID-19 cases among volunteers who received the AVX/COVID-12 booster compared to those who received the AZ/ChAdOx-1-S vaccine, supporting the conclusion of non-inferiority. Consequently, the AVX/COVID-12 vaccine meets the WHO’s non-inferiority standard when compared to AZ/ChAdOx-1-S. These compelling findings strongly advocate for the incorporation of the AVX/COVID-12 vaccine as a booster dose for the general population.

## Data Availability

The protocol was registered in the National Registry of Clinical Studies under number RNEC2022-AVXSARSCoV2VAC005 and published under NCT05710783. Individual de-identified participant data will not be shared beyond the limits permitted by the informed consent and Mexican law. Specifically, this includes the sharing of the study protocol, statistical analysis plan, informed consent form, and approved clinical study report. Additionally, other de-identified data allowed under the informed consent and Mexican law may be shared. The data will be made available immediately upon publication and for 12 months thereafter. Access to the data will be granted solely to investigators with methodologically sound proposals, subject to authorization by an independent review committee and the ethics committees involved in approving the protocol. If required by law, authorization from the Federal Commission for the Protection against Sanitary Risks (COFEPRIS) in Mexico will also be obtained. Any use of the data must strictly adhere to the authorized purposes outlined during the approval process.

## Acknowledgments

From CONAHCYT, we acknowledge María Elena Álvarez-Buylla Roces and Delia Aideé Orozco Hernández for their role as an inter-institutional liaison and overall facilitation of proceedings, committee evaluations, sanitary setup, and follow-up of trial design approvals, evaluations, and progress. We additionally acknowledge the broader support from various teams within all the clinical research sites and iLS Clinical Research S.C., particularly Gabriela Rosas, Daniela Castro, Claudia Aguilar, Roman Jarosch, and Alejandro Arias. From Laboratorio Avi-Mex, S. A. de C. V., we acknowledge the following people for their operative support: Bernardo Lozano Alcántara, Alejandro Ruiz, Rosalba Rodríguez, Leticia Espinosa Gervasio, Rodrigo Yebra Reyes, Vanessa Escamilla Jiménez, Jaime Becerra Jiménez, Lorena Juárez Pedraza, Sandra Yuridia Ang Tinajero, Avelia Ariadna Cuevas Cifuentes, Juan Pablo Robles Álvarez, Avirán Almazán Gutiérrez, Aurora Betsabé Gutiérrez Balderas, Merlenne Rubio Díaz, and Guadalupe Aguilar Rafael. From INER, we acknowledge the following people for their technical support: Liliana Figueroa Hernández, Francisco Cruz Flores, Claudia Ivett Hernández Lazaro, María Angelica Velázquez González, Jessica Romero Rodríguez, Dulce Cinthia Soriano Hernández, Milton Nieto Ponce, Edgar Reyna, Itzel Corona, José E. Marquez, Angelica Moncada, and Pablo Franco-Mendoza. From IMSS, we acknowledge the help of Diego Lozano-Cisneros in formatting the manuscript. From the Icahn School of Medicine at Mount Sinai, we acknowledge Sean Whelan. From Instituto Politécnico Nacional, Escuela Nacional de Ciencias Biológicas, we acknowledge Sonia Mayra Pérez Tapia, Alexis Gabriel Suárez Gómez, and Sandra Comparan Alarcón.

## Ethical Responsibilities

The protocol was approved by the ethics, research, and biosafety committees of each clinical research site, National Committee for Science, Technology and Innovation in Public Health of the National Council for Humanities, Science, and Technology (CONAHCYT), and also by the Federal Commission for the Protection against Sanitary Risks (COFEPRIS, Mexico) as a regulatory agency.

## Protection of Humans and Animals

No animals were used in this study. The protocol was designed to comply with the ethical principles of the Helsinki Declaration, Good Clinical Practices, and the applicable Mexican law.

## Privacy Rights and Informed Consent

Informed Consent formats were provided and signed by all participants under Good Clinical Practices and Mexican law. Personal data is confidential, and subjects are not identified personally. Their data was analyzed and duly de-identified.

---

**Supplementary Figure 1.**
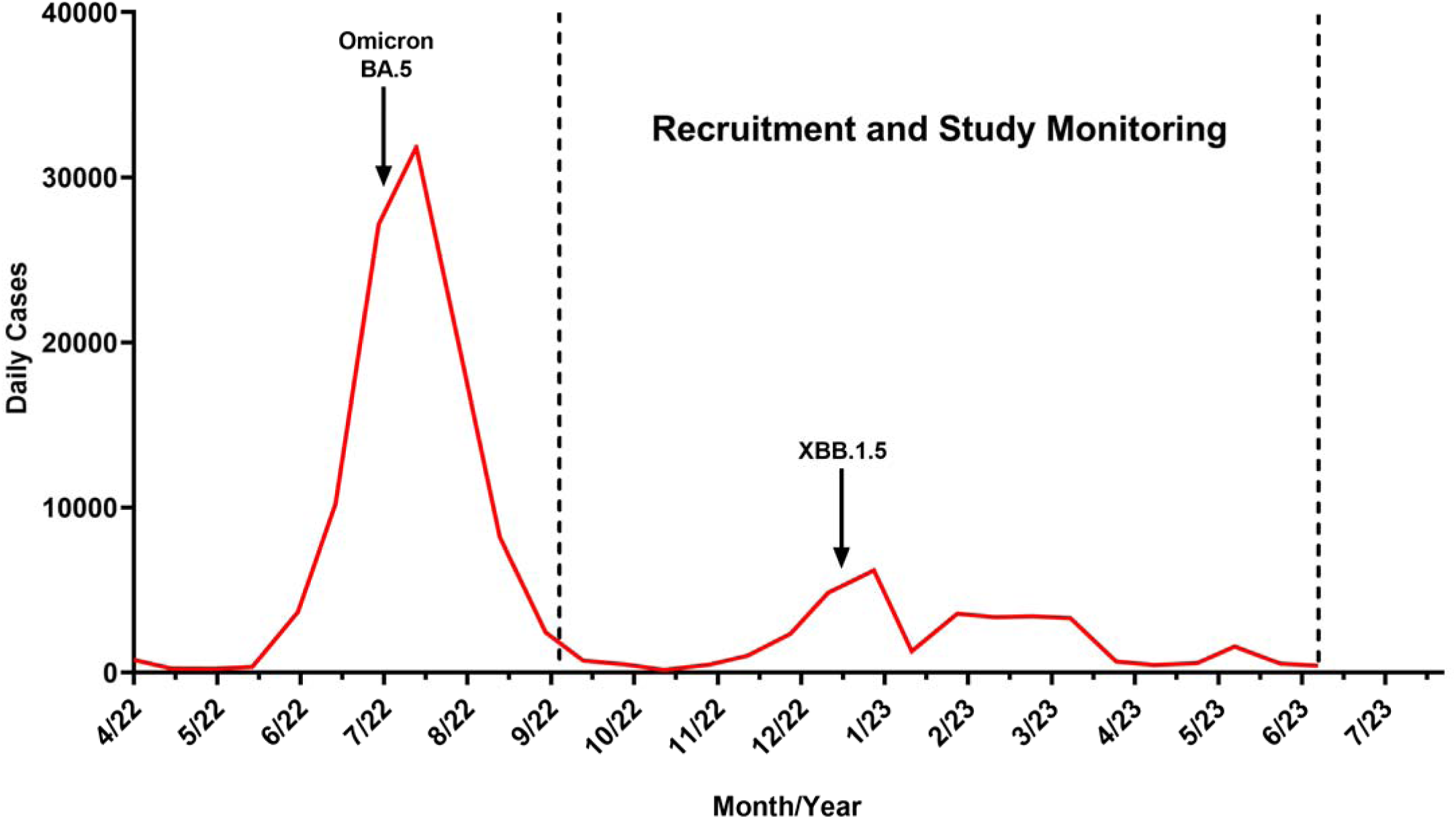
COVID-19 incidence in Mexico during the recruitment and monitoring of the study participants.

**Supplementary Figure 2.**
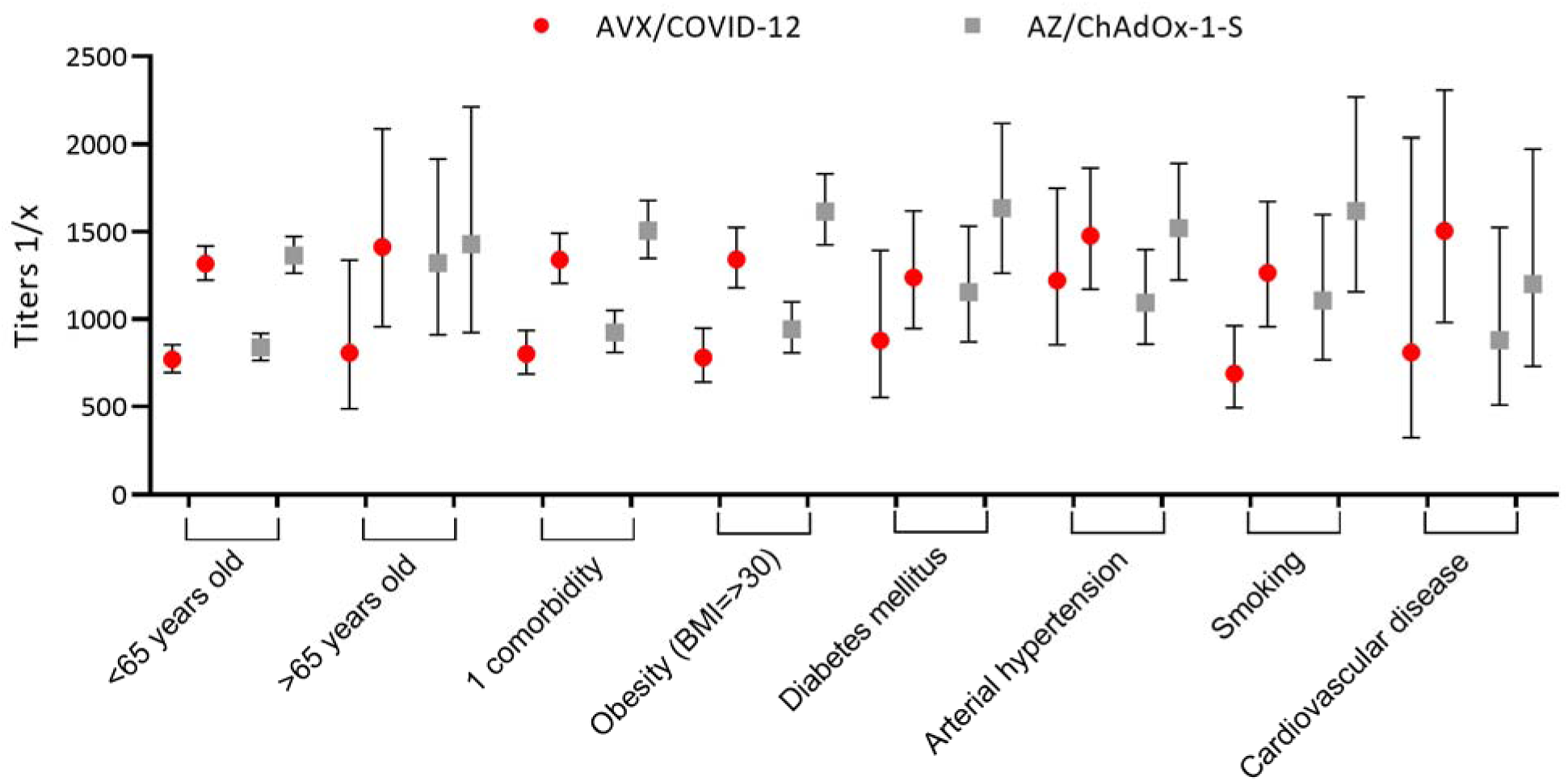
AVX/COVID-12 and AZ/ChAdOx-1-S Boosting Elicited Comparable Increases in Neutralizing Antibody Titers Among Participants with Comorbidities, as well as in Subjects >65 and <65 Years Old. The geometric mean and confidence interval for antibody titers at day 0 and day 14 are displayed for AVX/COVID-12 (red circles) and AZ/ChAdOx-1-S (gray squares). Both vaccines exhibited comparable rises in neutralizing antibody titers among participants with comorbidities and in those aged >65 and <65 years old.

**Supplementary Table 1.** Proportion of affected subjects by local adverse events of special interest at 7 days post-immunization.

| SOC | PT | Severity | AVX | % | AZ | % | Total | % | p-Value |
| --- | --- | --- | --- | --- | --- | --- | --- | --- | --- |
| Subjects |  |  | 3120 |  | 936 |  | 4056 |  |  |
| Any AE |  | Total | 800 | (25.64) | 193 | (20.61) | 993 | (24.48) | 0.001 |
|  |  | Mi | 698 | (22.37) | 173 | (18.48) | 871 | (21.47) | 0.01 |
|  |  | Mo | 108 | (3.46) | 21 | (2.24) | 129 | (3.18) | 0.06 |
|  |  | S | 9 | (0.28) | 0 | (0) | 9 | (0.22) | 0.10 |
| Skin and subcutaneous tissue disorders |  | Total | 23 | (0.73) | 5 | (0.53) | 28 | (0.69) | 0.51 |
|  |  | Mi | 17 | (0.54) | 5 | (0.53) | 22 | (0.54) | 0.96 |
|  |  | Mo | 6 | (0.19) | 0 | (0) | 6 | (0.14) | 0.17 |
|  |  | S | 0 | (0) | 0 | (0) | 0 | (0) |  |
|  | Erythema | Mo | 1 | (0.03) | 0 | (0) | 1 | (0.02) | 0.58 |
|  | Papule | Mi | 2 | (0.06) | 1 | (0.1) | 3 | (0.07) | 0.67 |
|  | Itching | Mi | 15 | (0.48) | 4 | (0.42) | 19 | (0.46) | 0.83 |
|  | Itching | Mo | 5 | (0.16) | 0 | (0) | 5 | (0.12) | 0.22 |
| Nervous system disorders |  | Total | 6 | (0.19) | 0 | (0) | 6 | (0.14) | 0.17 |
|  |  | Mi | 5 | (0.16) | 0 | (0) | 5 | (0.12) | 0.22 |
|  |  | Mo | 1 | (0.03) | 0 | (0) | 1 | (0.02) | 0.58 |
|  |  | S | 0 | (0) | 0 | (0) | 0 | (0) |  |
|  | Paresthesia | Mi | 5 | (0.16) | 0 | (0) | 5 | (0.12) | 0.22 |
|  | Paresthesia | Mo | 1 | (0.03) | 0 | (0) | 1 | (0.02) | 0.58 |
| General disorders and local events |  | Total | 786 | (25.19) | 190 | (20.29) | 976 | (24.06) | 0.002 |
|  |  | Mi | 685 | (21.95) | 170 | (18.16) | 855 | (21.07) | 0.01 |
|  |  | Mo | 101 | (3.23) | 21 | (2.24) | 122 | (3) | 0.11 |
|  |  | S | 9 | (0.28) | 0 | (0) | 9 | (0.22) | 0.10 |
|  | Heat at the vaccination site | Mi | 1 | (0.03) | 0 | (0) | 1 | (0.02) | 0.58 |
|  | Pain at the application site | Mi | 3 | (0.09) | 0 | (0) | 3 | (0.07) | 0.34 |
|  | Pain at the application site | Mi | 655 | (20.99) | 165 | (17.62) | 820 | (20.21) | 0.02 |
|  | Pain at the injection site | Mo | 98 | (3.14) | 21 | (2.24) | 119 | (2.93) | 0.15 |
|  | Pain at the injection site | S | 8 | (0.25) | 0 | (0) | 8 | (0.19) | 0.12 |
|  | Edema at the injection site | Mi | 4 | (0.12) | 1 | (0.1) | 5 | (0.12) | 0.87 |
|  | Edema at the injection site | Mo | 5 | (0.16) | 0 | (0) | 5 | (0.12) | 0.22 |
|  | Redness at the injection site | Mi | 28 | (0.89) | 9 | (0.96) | 37 | (0.91) | 0.85 |
|  | Redness at the injection site | Mo | 7 | (0.22) | 0 | (0) | 7 | (0.17) | 0.14 |
|  | Redness at the injection site | S | 1 | (0.03) | 0 | (0) | 1 | (0.02) | 0.58 |
|  | Hematoma at the injection site | Mi | 1 | (0.03) | 1 | (0.1) | 2 | (0.04) | 0.36 |
|  | Swelling | Mi | 4 | (0.12) | 2 | (0.21) | 6 | (0.14) | 0.55 |
|  | Induration at the application site | Mi | 11 | (0.35) | 3 | (0.32) | 14 | (0.34) | 0.88 |
|  | Inflammation | Mi | 13 | (0.41) | 1 | (0.1) | 14 | (0.34) | 0.15 |
|  | Nerve injury at the injection site | Mi | 0 | (0) | 2 | (0.21) | 2 | (0.04) | 0.009 |
|  | Itching at the injection site | Mi | 0 | (0) | 1 | (0.1) | 1 | (0.02) | 0.07 |
| Musculoskeletal and connective tissue disorders |  | **Total** | 10 | (0.32) | 0 | (0) | 10 | (0.24) | 0.08 |
|  |  | **Mi** | 6 | (0.19) | 0 | (0) | 6 | (0.14) | 0.17 |
|  |  | **Mo** | 4 | (0.12) | 0 | (0) | 4 | (0.09) | 0.27 |
|  |  | **S** | 0 | (0) | 0 | (0) | 0 | (0) |  |
|  | Pain in a limb | Mi | 4 | (0.12) | 0 | (0) | 4 | (0.09) | 0.27 |
|  | Pain in a limb | Mo | 3 | (0.09) | 0 | (0) | 3 | (0.07) | 0.34 |
|  | Discomfort in the extremities | Mi | 2 | (0.06) | 0 | (0) | 2 | (0.04) | 0.43 |
|  | Discomfort in the extremities | Mo | 1 | (0.03) | 0 | (0) | 1 | (0.02) | 0.58 |

SOC: System Organ Class; PT: Preferred term; AE: Adverse Event; Mi: Mild; Mo: Moderate; S: Severe; AVX: AVX/COVID-12; AZ: AZ/ChAdOx-1-S. P-value calculated from comparison of proportions (Z-test)

**Supplementary Table 2.**
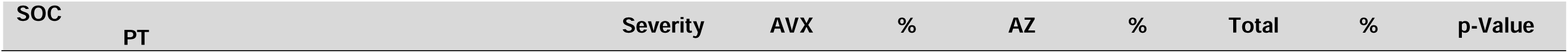
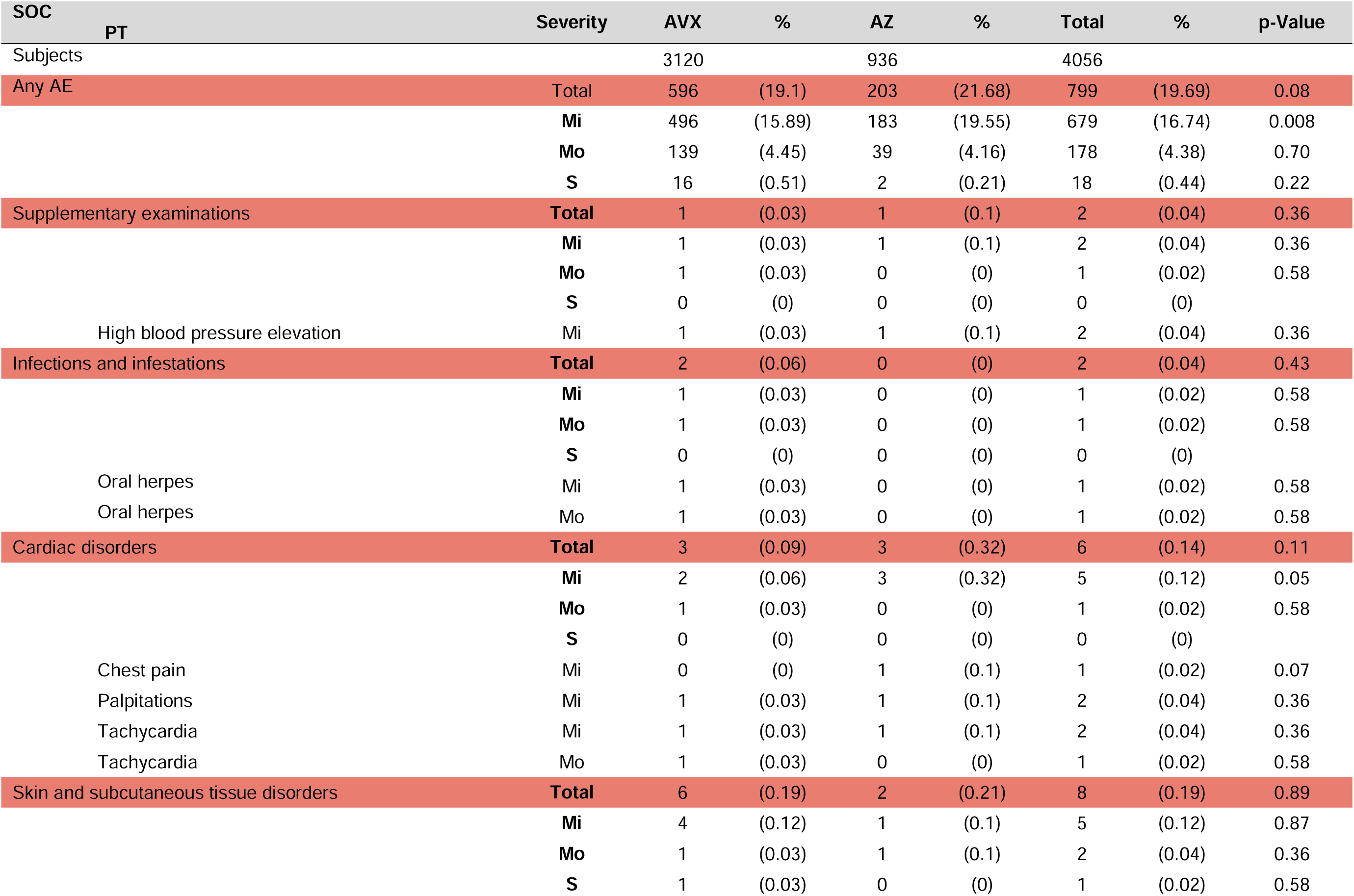
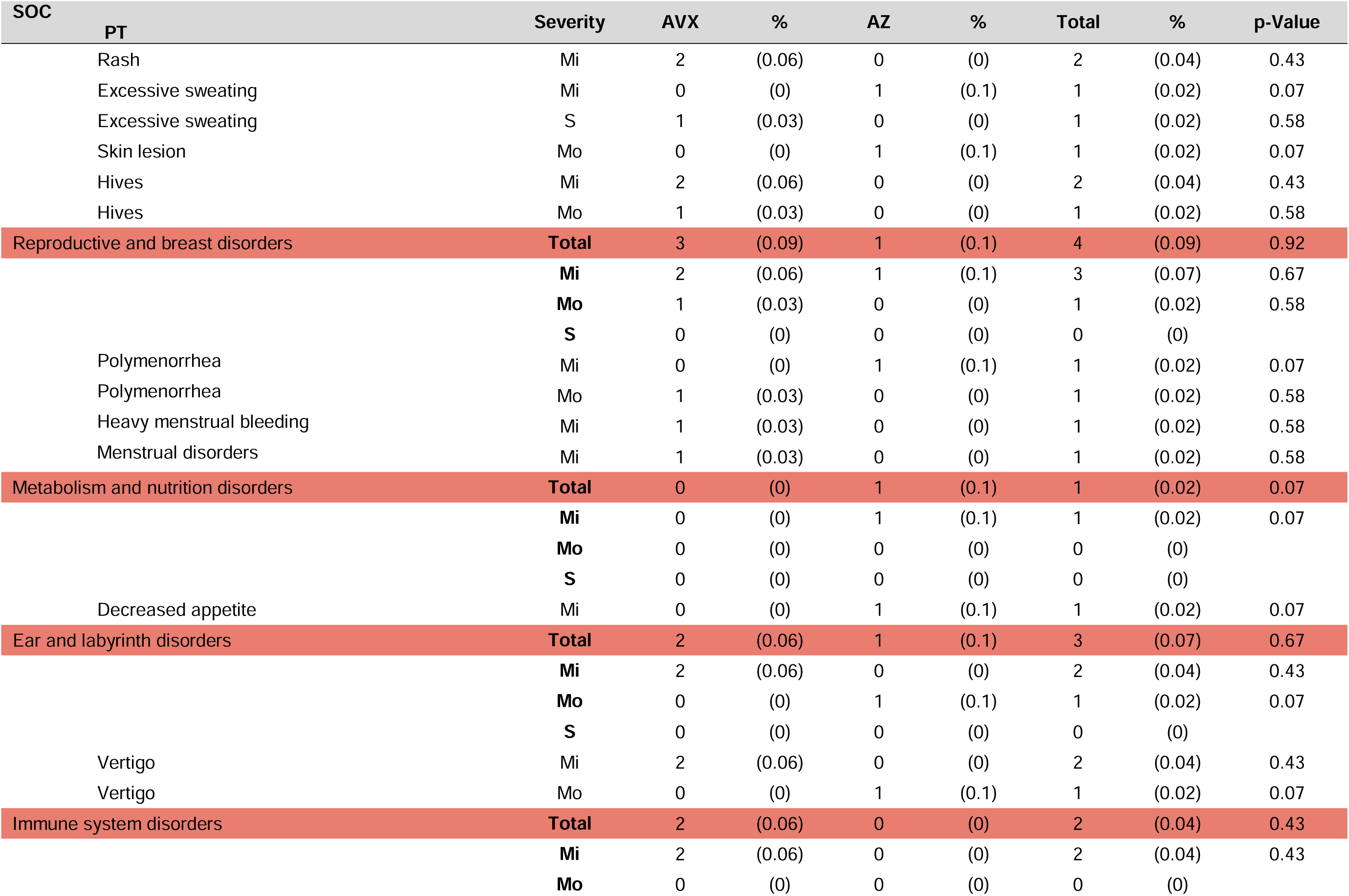
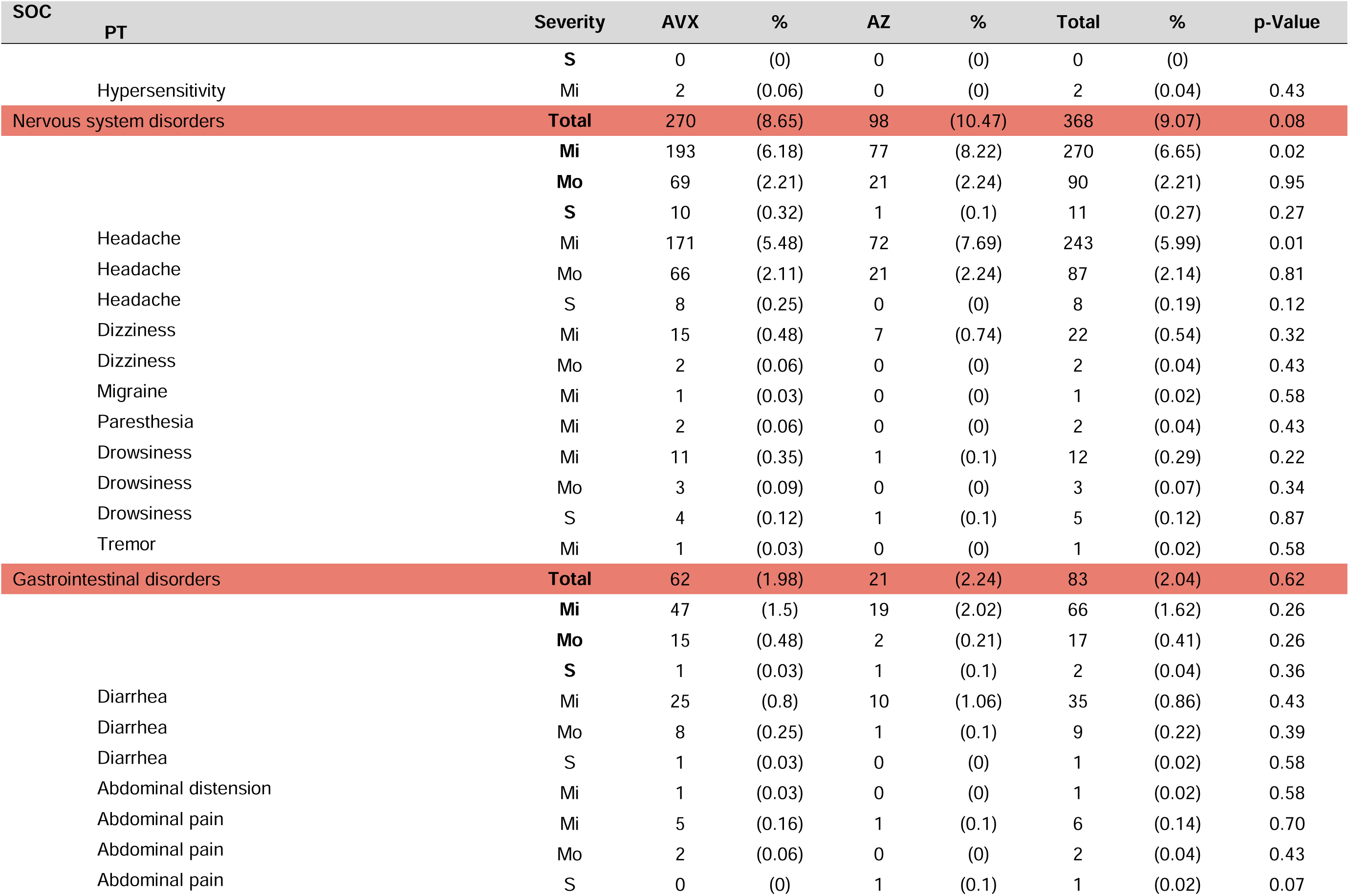
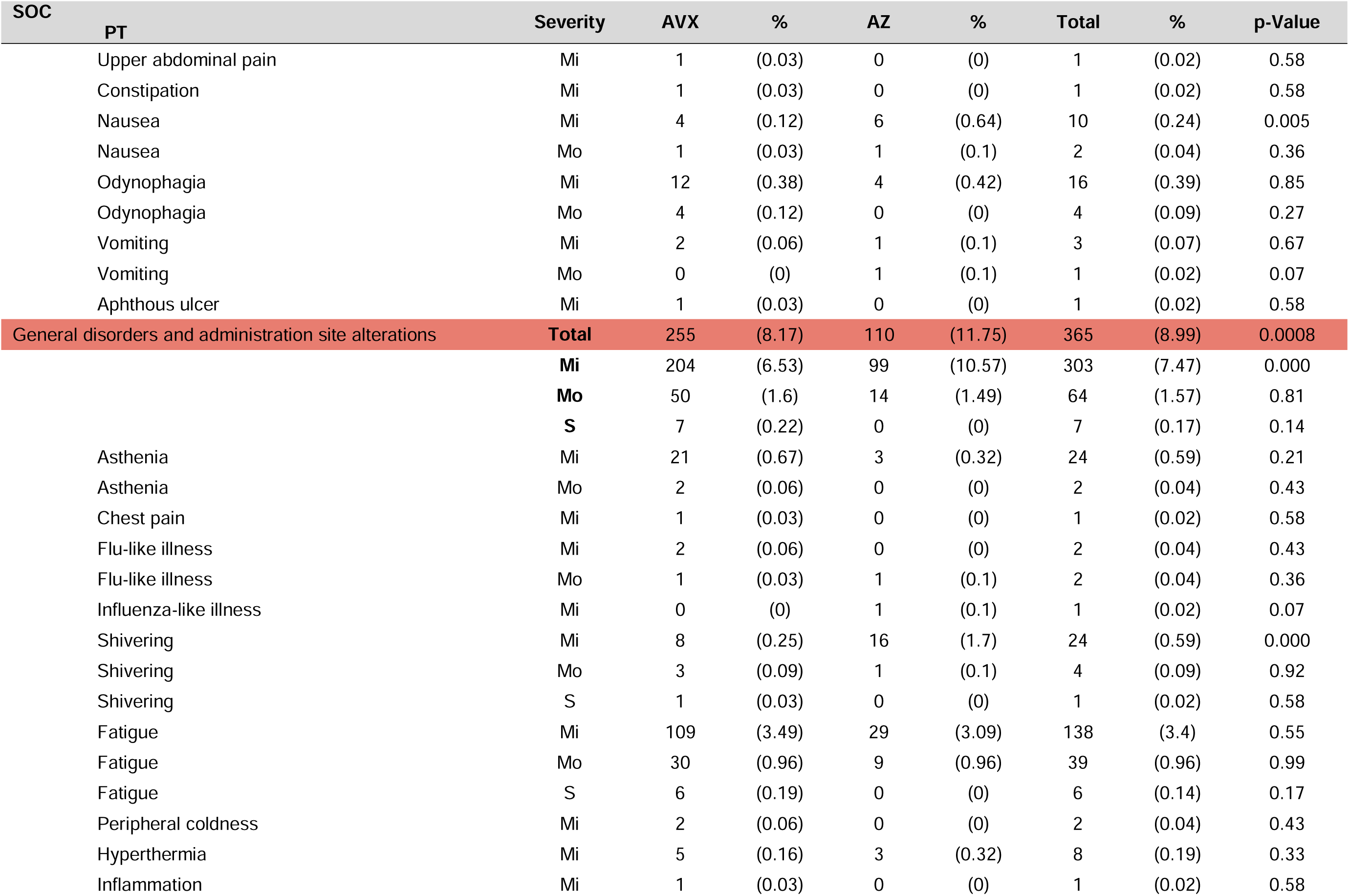
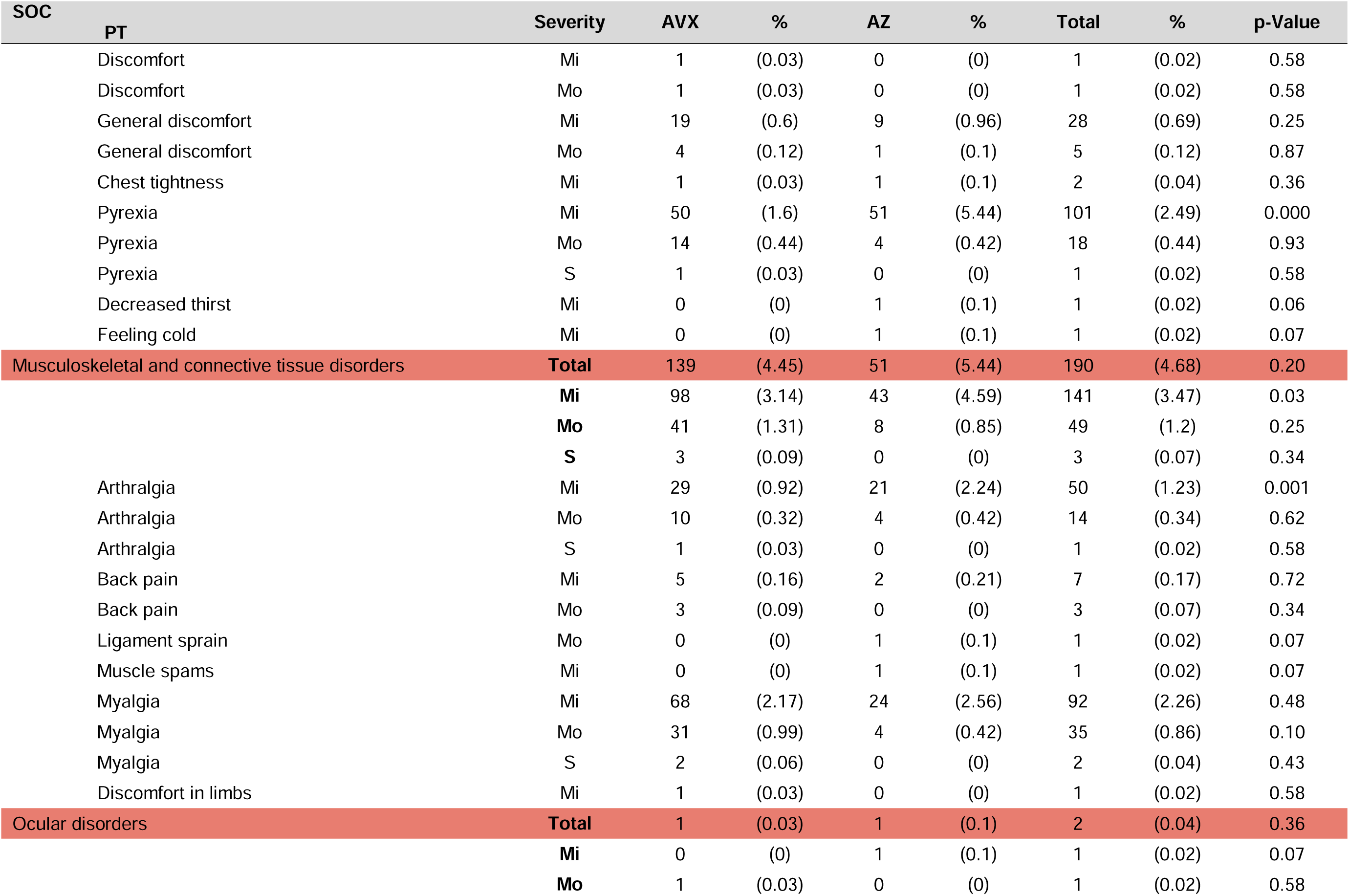
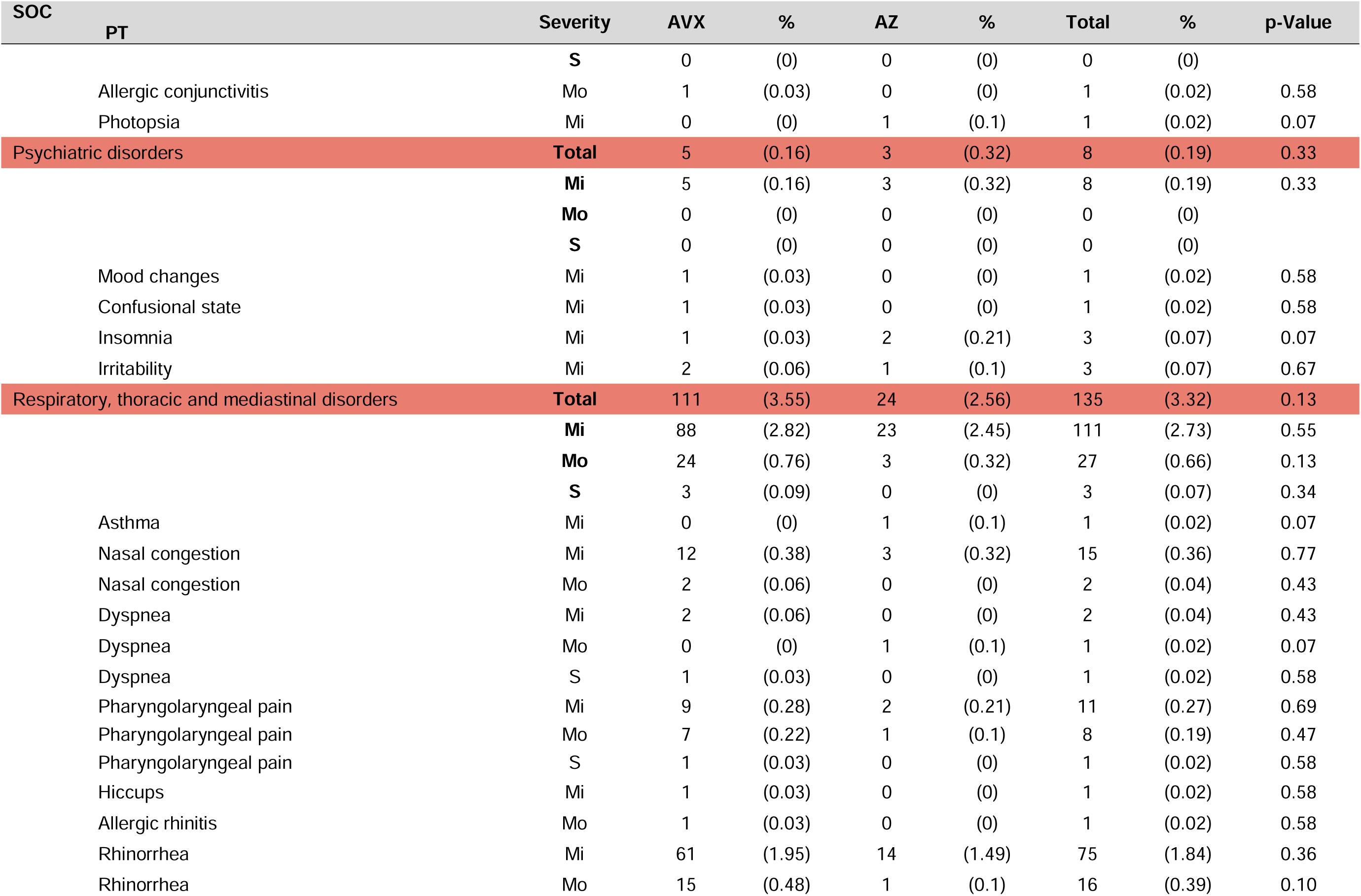
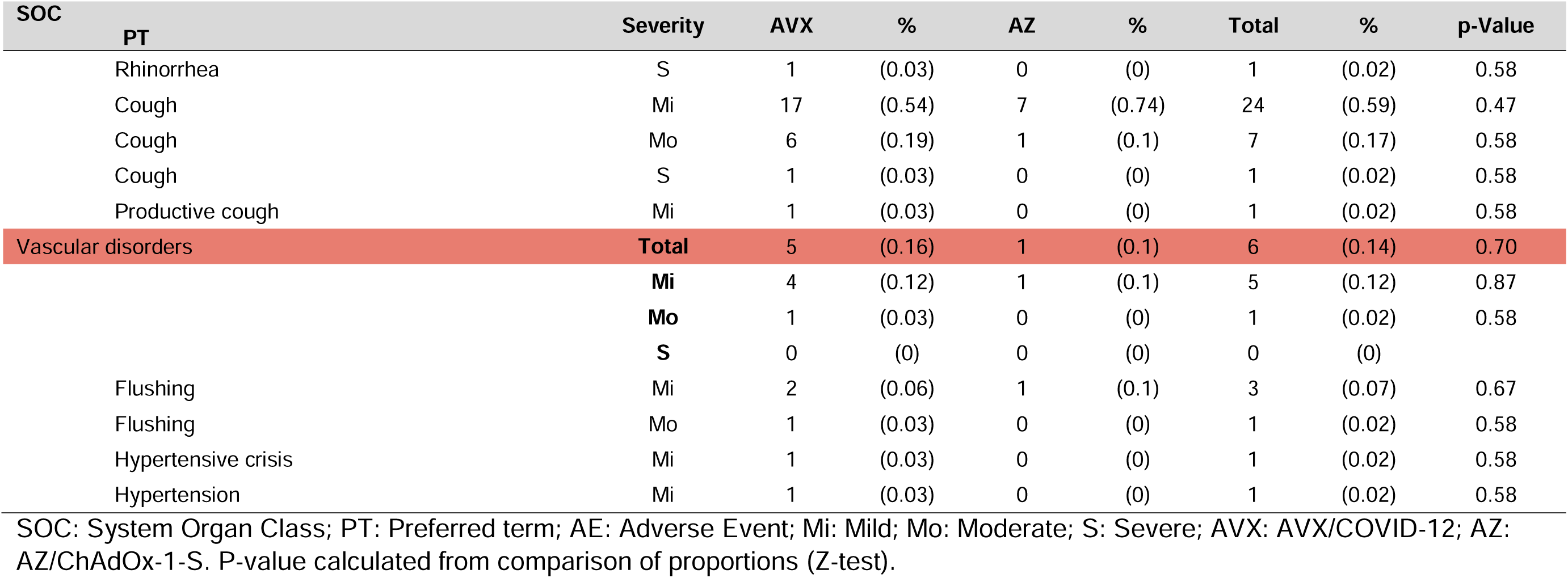
Proportion of affected subjects by systemic adverse events of special interest at 7 days post-immunization.

**Supplementary Table 3.**
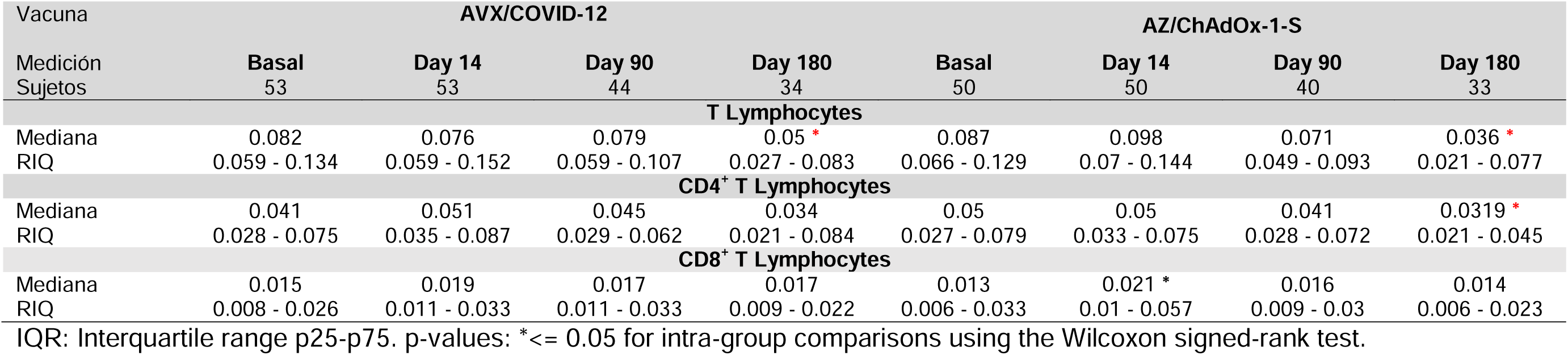
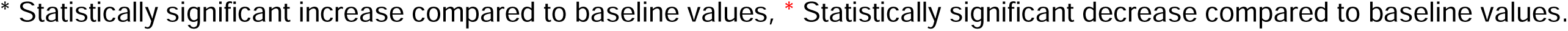
Percentage of total, CD4^+^ and CD8^+^ T cells producing IFN-γ in response to fraction 1 of spike protein stimulation.

| Vacuna |  | AVX/COVID-12 |  |  |  | AZ/ChAdOx-1-S |  |  |  |
| --- | --- | --- | --- | --- | --- | --- | --- | --- | --- |
| Medición |  | Basal | Day 14 | Day 90 | Day 180 | Basal | Day 14 | Day 90 | Day 180 |
| Sujetos |  | 53 | 53 | 44 | 34 | 50 | 50 | 40 | 33 |
| **T Lymphocytes** |  |  |  |  |  |  |  |  |  |
| Mediana |  | 0.082 | 0.076 | 0.079 | 0.05 * | 0.087 | 0.098 | 0.071 | 0.036 * |
| RIQ |  | 0.059 - 0.134 | 0.059 - 0.152 | 0.059 - 0.107 | 0.027 - 0.083 | 0.066 - 0.129 | 0.07 - 0.144 | 0.049 - 0.093 | 0.021 - 0.077 |
| **CD4<sup>+</sup> T Lymphocytes** |  |  |  |  |  |  |  |  |  |
| Mediana |  | 0.041 | 0.051 | 0.045 | 0.034 | 0.05 | 0.05 | 0.041 | 0.0319 * |
| RIQ |  | 0.028 - 0.075 | 0.035 - 0.087 | 0.029 - 0.062 | 0.021 - 0.084 | 0.027 - 0.079 | 0.033 - 0.075 | 0.028 - 0.072 | 0.021 - 0.045 |
| **CD8<sup>+</sup> T Lymphocytes** |  |  |  |  |  |  |  |  |  |
| Mediana |  | 0.015 | 0.019 | 0.017 | 0.017 | 0.013 | 0.021 * | 0.016 | 0.014 |
| RIQ |  | 0.008 - 0.026 | 0.011 - 0.033 | 0.011 - 0.033 | 0.009 - 0.022 | 0.006 - 0.033 | 0.01 - 0.057 | 0.009 - 0.03 | 0.006 - 0.023 |

IQR: Interquartile range p25-p75. p-values: \*≤ 0.05 for intra-group comparisons using the Wilcoxon signed-rank test.
\* Statistically significant increase compared to baseline values, \* Statistically significant decrease compared to baseline values.

